# The effect of anti-SARS-CoV-2 monoclonal antibody, bamlanivimab, on endogenous immune response to COVID-19 vaccination

**DOI:** 10.1101/2021.12.15.21267605

**Authors:** Robert J. Benschop, Jay L. Tuttle, Lin Zhang, Josh Poorbaugh, Nicole L. Kallewaard, Peter Vaillancourt, Melissa Crisp, Thi Ngoc Vy Trinh, Joshua J. Freitas, Stephanie Beasley, Montanea Daniels, Natalie Haustrup, Richard E. Higgs, Ajay Nirula, Myron S. Cohen, Mary Marovich

## Abstract

As the COVID-19 pandemic evolves, and vaccine rollout progresses, the availability and demand for monoclonal antibodies for the prevention and treatment of SARS-CoV-2 infection are also accelerating. This longitudinal serological study evaluated the magnitude and potency of the endogenous antibody response to COVID-19 vaccination in participants who first received a COVID-19 monoclonal antibody in a prevention study. Over the course of six months, serum samples were collected from the prevention population (nursing home residents and staff) enrolled in the BLAZE-2 clinical trial who had received either bamlanivimab (4200 mg) or placebo. In an unplanned component of this trial, a subset of these participants was subsequently fully vaccinated with two doses of either SpikeVax (Moderna) or Comirnaty (BioNTech/Pfizer) COVID-19 mRNA vaccines, as part of the US vaccination program. This post-hoc analysis assessed the immune response to vaccination for the subset of participants (N=135) without prior SARS-CoV-2 infection. Antibody titers and potency were assessed using three assays against SARS-CoV-2 proteins that bamlanivimab does not significantly bind to, thereby reflecting the endogenous antibody response. All bamlanivimab and placebo participants mounted a robust immune response to full COVID-19 vaccination, irrespective of age, risk-category and vaccine type, with any observed differences unlikely to be clinically meaningful. These findings are pertinent for informing public health policy with results that suggest a complementary role for COVID-19 monoclonal antibodies (mAbs) with COVID-19 vaccines and that the benefit of receiving COVID-19 vaccination at the earliest opportunity outweighs the minimal effect on the endogenous immune response due to prior prophylactic COVID-19 mAb infusion.

**One Sentence Summary:** Individuals infused with an anti-SARS-CoV-2 antibody demonstrated a robust immune response to subsequent full COVID-19 vaccination.

## Introduction

In response to the coronavirus disease 2019 (COVID-19) pandemic, many prophylactic and therapeutic treatments were rapidly developed to target the highly pathogenic severe acute respiratory syndrome coronavirus 2 (SARS-CoV-2) *(1, 2)*. Antibodies that target the Receptor Binding Domain (RBD) of the spike protein of SARS-CoV-2 are essential for protection against COVID-19 disease *(3, 4)*, as these antibodies reduce SARS-CoV-2 viral load, which is correlated with disease severity *(5-7)*. Active immunity against COVID-19 disease develops when endogenous RBD-neutralizing antibodies are elicited following exposure to a pathogenic agent, such as SARS-CoV-2 or a COVID-19 vaccine *(3, 4)*; and passive immunity is conferred through the administration of exogenous antibodies, such as mAbs, that precisely target and bind to the RBD. Several authorized clinically active COVID-19 mAbs provide immediate protective immunity that persists for as long as the antibody concentration exceeds that required for neutralization of the virus *(8-12)*. Bamlanivimab was the first COVID-19 mAb to be granted Emergency Use Authorization (EUA) in November 2020 by the U.S. Food and Drug Administration (FDA), but was later revoked in April 2021, due to the increase of SARS-CoV-2 viral variants that were resistant to bamlanivimab alone *(11, 13)*. Vaccine-induced protection develops over time, often requiring multiple doses but offers clear advantages by eliciting a broader polyclonal immune response and establishing immunological memory for durable immunity *(14, 15)*.

There are currently insufficient safety and efficacy data with COVID-19 vaccines in individuals who have previously received COVID-19 mAbs; in the absence of data both the World Health Organization (WHO) and the Centers for Disease Control and Prevention (CDC) recommend the deferral of vaccination for 90 days following mAb treatment and more recently, if mAbs were received for post-exposure prophylaxis (PEP), CDC now recommends vaccine deferral for 30 days *(16, 17)*. In order to avoid unnecessary delays for individuals seeking vaccination and to inform public health policy, it is critical that we understand any effect that therapeutic mAbs have on the subsequent vaccine-induced immune response.

Vaccine efficacy against COVID-19 disease is correlated with the elicitation of antibodies, and accordingly serological assays are critical tools for monitoring the longitudinal endogenous antibody responses following COVID-19 treatments or SARS-CoV-2 infection *(18-20)*. In a prospective treatment case study, an individual who was treated with an anti-SARS-CoV-2 mAb for symptomatic COVID-19 and received mRNA COVID-19 vaccination more than 40 days thereafter, exhibited comparable post-vaccine antibody responses to SARS-CoV-2 RBD for SARS-CoV-2 variants (including Alpha, Beta and Gamma), to other participants who had not received an anti-SARS-CoV-2 mAb and were vaccinated following COVID-19 *(21)*. However, larger studies are required to assess the duration of exogenous anti-SARS-CoV-2 mAbs in individuals with COVID-19 disease and whether these mAbs interfere with a subsequent immune response to a later COVID-19 vaccine. Additional studies are also required to assess the potential impact of prophylactic treatment with anti-SARS-CoV-2 mAbs on the specificity, magnitude, functionality, and duration of the endogenous antibody response to SARS-CoV-2 infection or COVID-19 vaccination. Here we study the singular question of whether the presence of prophylactic mAbs in SARS-CoV-2-naïve individuals interfere with endogenous immune responses to vaccination. We present the results from a post-hoc analysis of immune responses to full COVID-19 vaccination with either SpikeVax (mRNA-1273, Moderna) or Comirnaty (BNT162b2, BioNTech/Pfizer) mRNA vaccines following passive immunization with bamlanivimab mAb administered as a COVID-19 prevention intervention for participants who were residents or staff of US skilled nursing and assisted living facilities *(22)*.

## Results

### Participant Characteristics

The BLAZE-2 (NCT04497987) clinical trial was a phase 3, randomized, double-blind, placebo-controlled, single-dose study to evaluate whether bamlanivimab prevents SARS-CoV-2 infection in staff and residents of skilled nursing and assisted living facilities with a high risk of SARS-CoV-2 exposure. This post-hoc analysis included a total of 499 samples from 135 SARS-CoV-2-naïve participants who received either bamlanivimab (4,200 mg) or placebo (day 1) during the BLAZE-2 prophylaxis study and were subsequently fully vaccinated within the scheduled serum sampling period of the trial (day 169). The CDC describes an individual as fully vaccinated 2 weeks after the second COVID-19 vaccine dose in a 2-dose series, such as for Comirnaty or SpikeVax *(23)*.

There is a linear relationship between bamlanivimab dose and exposure as determined in an earlier pharmacokinetics modelling study and the half-life of bamlanivimab is approximately 17 days *(24)*. Participants received the first COVID-19 vaccine dose at different timepoints (ranging from 43 to 127 days, median 67 days) following bamlanivimab or placebo infusion. A total of 95 participants (70%) received the first vaccine dose within 90 days of the bamlanivimab or placebo infusion. Most participants received the second COVID-19 vaccine dose following the recommended interval specified in the EUA factsheet for each vaccine (21 and 28 days later for Comirnaty and SpikeVax, respectively) *(25, 26)*. A total of 96 participants (71%) received the Comirnaty vaccine and 39 participants (29%) received the SpikeVax vaccine.

An overview of the post-hoc analysis and participant inclusion criteria for each analysis are detailed in the Materials and Methods section. The baseline characteristics of the participants included in this analysis are shown in Table 1.

**Table 1:** Baseline characteristics of participants (N=135) included in post-hoc analysis

| Characteristics |  | Bamlanivimab (4,200 mg) |  | Placebo |  |
| --- | --- | --- | --- | --- | --- |
|  |  | Residents | Staff | Residents | Staff |
|  |  | N=22 | N=51 | N=14 | N=48 |
| Age, | Median | 63 | 43 | 82 | 44 |
|  | (range) | (31 - 95) | (20 - 74) | (63 - 93) | (19 - 67) |
| Sex, | No. (%) |  |  |  |  |
|  | Male | 10 (45%) | 11 (22%) | 5(36%) | 4 (8%) |
|  | Female | 12 (55%) | 40 (78%) | 9 (64%) | 44 (92%) |
| High-risk of severe COVID-19 disease, |  |  |  |  |  |
|  | No. (%) | 22 (100%) | 24 (47%) | 14 (100%) | 21 (44%) |

*High-risk categorization criteria (13)*

The median age of staff participants (N=99) was 43 years compared with resident participants (N=36) who had a median age of 72 years. Of the 99 staff participants, 6% were 65 years or older. A total of 81 participants (60%) met the criteria for high-risk of developing severe COVID-19 disease, which included 100% of the residents and 45% of the staff. The criteria for classifying high-risk of severe COVID-19 disease for this post-hoc analysis have been described previously *(13)*.

### Robust endogenous antibody response to full COVID-19 vaccination following anti-SARS-CoV-2 mAb infusion

A multiplex custom assay was performed on serum samples obtained from fully vaccinated participants (N=135) to measure the magnitude of the binding antibody response. The assay was performed against the spike-RBD carrying the E484Q alteration (spike-RBD-E484Q) and the spike N-terminal domain (spike-NTD) (Table 2). Since the epitope for bamlanivimab lies within the spike RBD *(27, 28)* and bamlanivimab does not significantly bind to the RBD with alterations at residue E484 *(29, 30)* or to NTD, antibody titers against these two SARS-CoV-2 proteins reflect the endogenous antibody response. Compared with placebo, treatment with bamlanivimab resulted in a 1.8-fold (p=0.001) and 2.0-fold (p<0.001) lower titer against spike-RBD-E484Q and spike-NTD, respectively (least square means comparison) (Fig. 1).

**Table 2:**
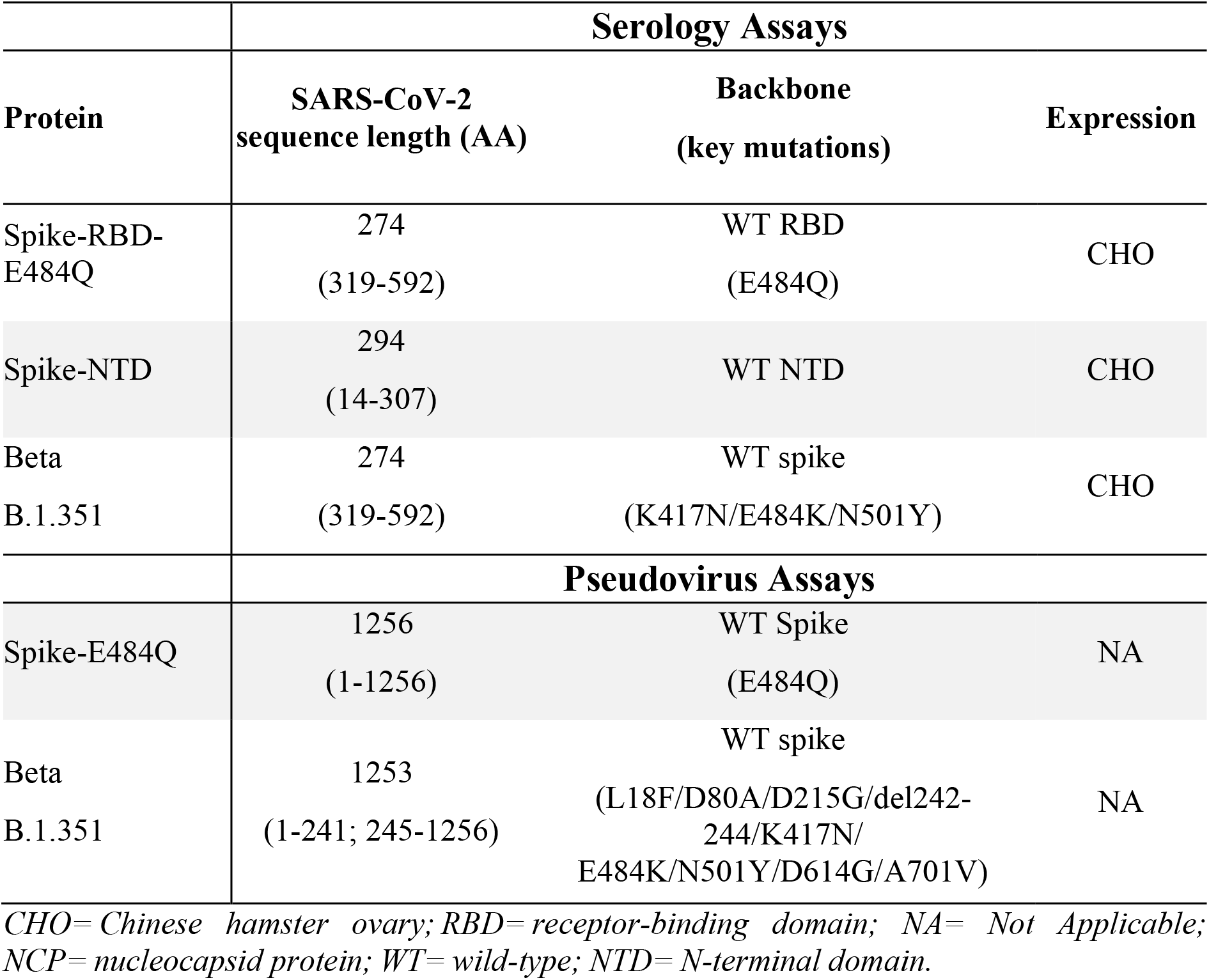
Details of SARS-CoV-2 proteins

**Fig. 1:**
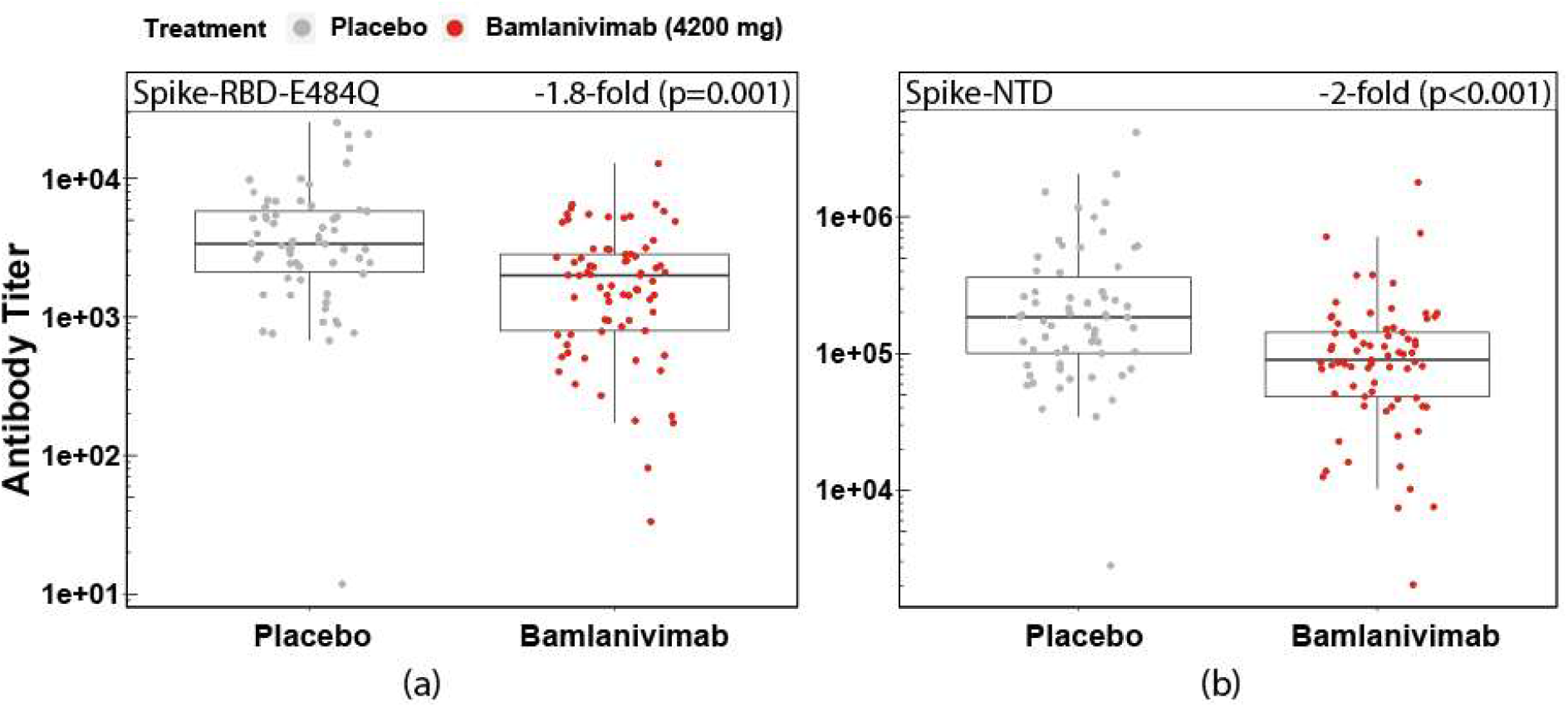
Binding antibody titers against (a) Spike-RBD-E484Q and (b) Spike-NTD for participants who had received either bamlanivimab or placebo infusion and were subsequently fully vaccinated (SpikeVax or Comirnaty) against COVID-19. Antibody titers were rescaled after adjusting for covariates. Boxes and horizontal bars denote the interquartile range (IQR) and the median, respectively. Length of whiskers corresponds to 1.5 times the IQR.

These binding antibody titer data were grouped into participants who were either staff or residents (Fig. 2a) and participants who received either SpikeVax or Comirnaty (Fig. 2b), to ascertain firstly, whether the immune response to full vaccination differs between these two groups and secondly, whether bamlanivimab infusion disparately affected these groups. Antibody titers from staff (median age 43 years) were 2.7 and 2.3 times (p<0.001) higher than titers from residents (median age 72 years) against Spike-RBD-E484Q and spike-NTD, respectively (Fig 2a). The effect of bamlanivimab on vaccine-induced antibody titer against Spike-RBD-E484Q and spike-NTD (p=0.388 and p=0.105, respectively) was similar for both residents and staff (Fig. 2a). There was no significant difference between antibody titers for participants who received SpikeVax or Comirnaty against either Spike-RBD-E484Q or spike-NTD (p=0.722 and p=0.397, respectively) (Fig. 2b). For participants who received either vaccine, the effect of bamlanivimab on the vaccine-induced antibody titer against Spike-RBD-E484Q and spike-NTD was also not significantly different (p=0.922 and p=0.756, respectively). These same trends were also observed for antibody titers measured against the SARS-CoV-2 beta variant (B.1.351), which is not recognized by bamlanivimab (fig. S1).

**Fig. 2:**
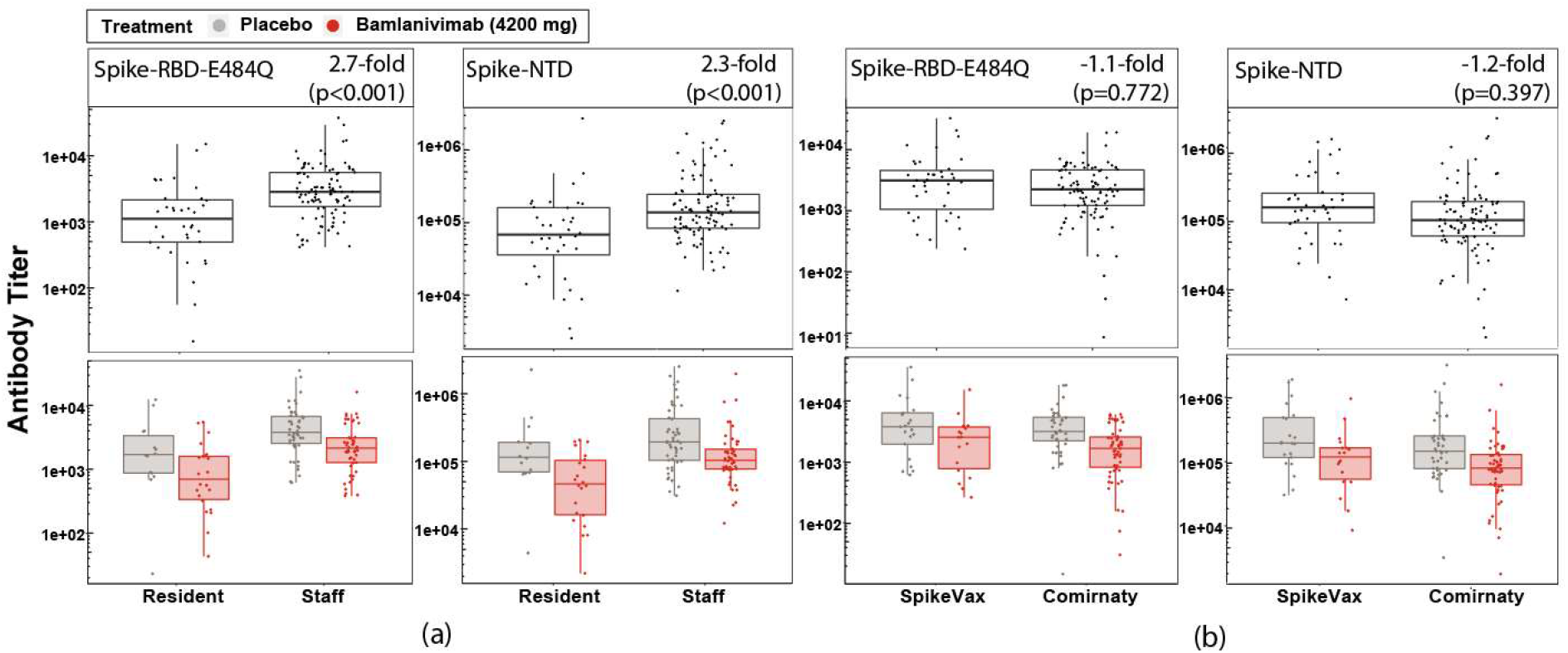
Binding antibody titers against Spike-RBD-E484Q and spike-NTD comparing samples from fully vaccinated participants who were (a) staff or resident (top) and further grouped by those who received placebo or bamlanivimab prior to vaccination (bottom) (b) participants who received SpikeVax or Comirnaty vaccine (top) and further grouped by those who received placebo or bamlanivimab prior to vaccination (bottom). Antibody titers were rescaled after adjusting for covariates. Boxes and horizontal bars denote the interquartile range (IQR) and the median, respectively. Length of whiskers corresponds to 1.5 times the IQR.

Since the staff (N=99) included participants who were at high-risk of developing severe COVID-19 disease, antibody titers were also compared between high-risk staff (N=45) with non-high-risk staff (N=54) (fig. S2). For high-risk staff compared with non-high-risk staff, the effect of bamlanivimab on the vaccine-induced antibody titer was similar (p=0.249) against spike-NTD and significantly lower (1.8-fold, p=0.037) against Spike-RBD-E484Q (fig. S2).

The potency of the endogenous antibodies produced in response to full vaccination was evaluated in two ways: based on angiotensin-converting enzyme 2 (ACE2) binding inhibition measured with the custom multiplex assay and pseudovirus neutralization activity using a vesicular stomatitis (VSV)-based pseudovirus. The ACE2 binding inhibition potency expresses how effectively the endogenous antibodies inhibited RBD-(E484Q)-ACE2 binding. Compared with placebo, receipt of bamlanivimab resulted in a 4.1-fold (p<0.001) lowering in ability of the endogenous antibody response to inhibit ACE2 binding (Fig. 3a). These ACE2 binding inhibition potency data were grouped into participants who were either staff or residents (Fig. 3b) and participants who received either SpikeVax or Comirnaty (Fig. 3c), to ascertain firstly, whether the potency of antibodies elicited following full vaccination differs between these groups and secondly, whether bamlanivimab infusion prior to full vaccination disparately affects these groups.

**Fig. 3:**
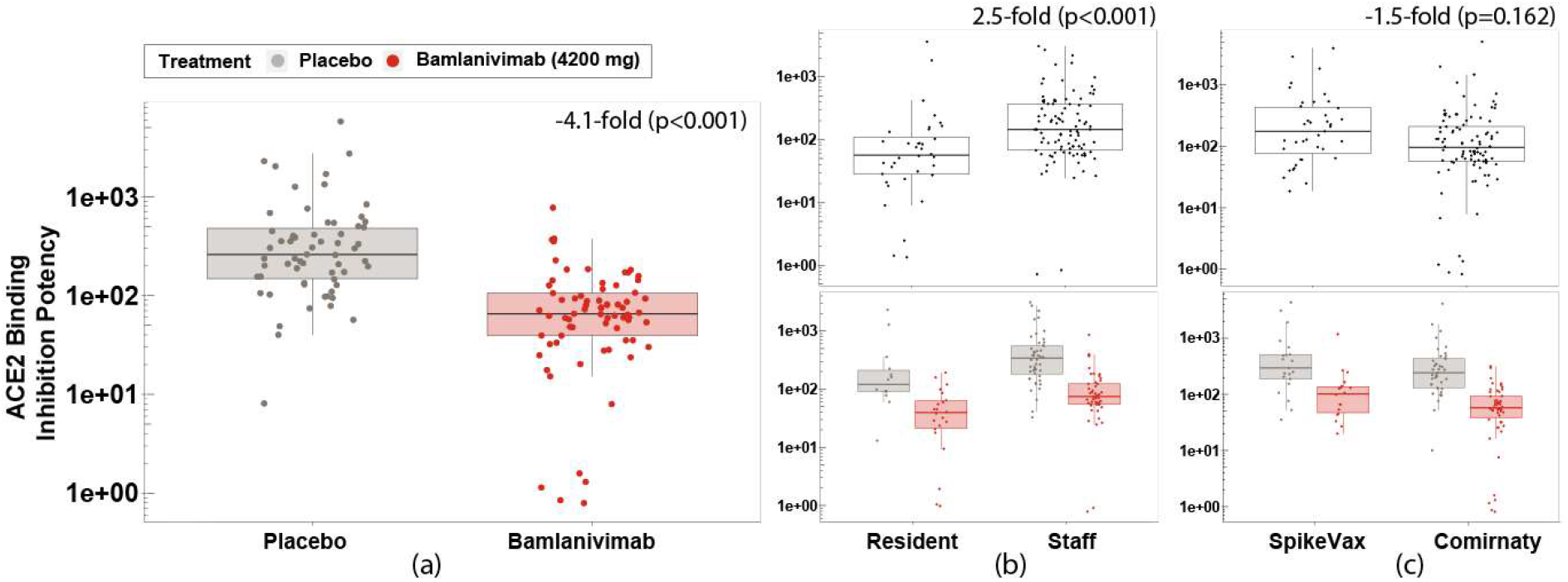
ACE2 binding inhibition potency (1/IC50) of serum samples collected from fully vaccinated participants (a) who had received placebo or bamlanivimab prior to vaccination (b) who were staff or resident (top) and further grouped by those who received placebo or bamlanivimab (bottom) (c) who received Comirnaty or SpikeVax (top) and further grouped by those who received placebo or bamlanivimab (bottom). ACE2 binding inhibition potency measured as 1/IC50 and adjusted for covariates. Boxes and horizontal bars denote the interquartile range (IQR) and the median reciprocal IC50, respectively. Length of whiskers corresponds to 1.5 times the IQR.

The ACE2 binding inhibition potency of endogenous antibodies was measured as 2.5 times (p<0.001) higher for staff than for residents (Fig. 3b). There was no disparity in ACE2 binding inhibition potency between participants who received SpikeVax or Comirnaty (p=0.162) (Fig. 3c). The magnitude of the bamlanivimab effect on ACE2 binding inhibition potency was similar for both resident and staff (p=0.233) (Fig. 3b) and for participants who had SpikeVax or Comirnaty (p=0.574) (Fig. 3c). We observed no difference in ACE2 binding inhibition potency between non-high-risk and high-risk staff (p=0.441), nor was the magnitude of the bamlanivimab effect on ACE2 binding inhibition potency different for these groups (p=0.084) (fig. S3a).

To assess the functional polyclonal antibody response against the full-length spike, neutralization potency was measured using a VSV-based pseudovirus for samples from a subset of participants (N=49; 21 placebo, 28 bamlanivimab). The participants sampled had all received their first vaccine dose 64 days or fewer (median 57 days) after a bamlanivimab or placebo infusion and thereby were the most likely to exhibit an effect of bamlanivimab infusion on the immune response to subsequent vaccination. There was no statistically significant difference in pseudovirus neutralization potency against Spike-E484Q for participants who received either placebo or bamlanivimab (p=0.078) (Fig. 4).

**Fig. 4:**
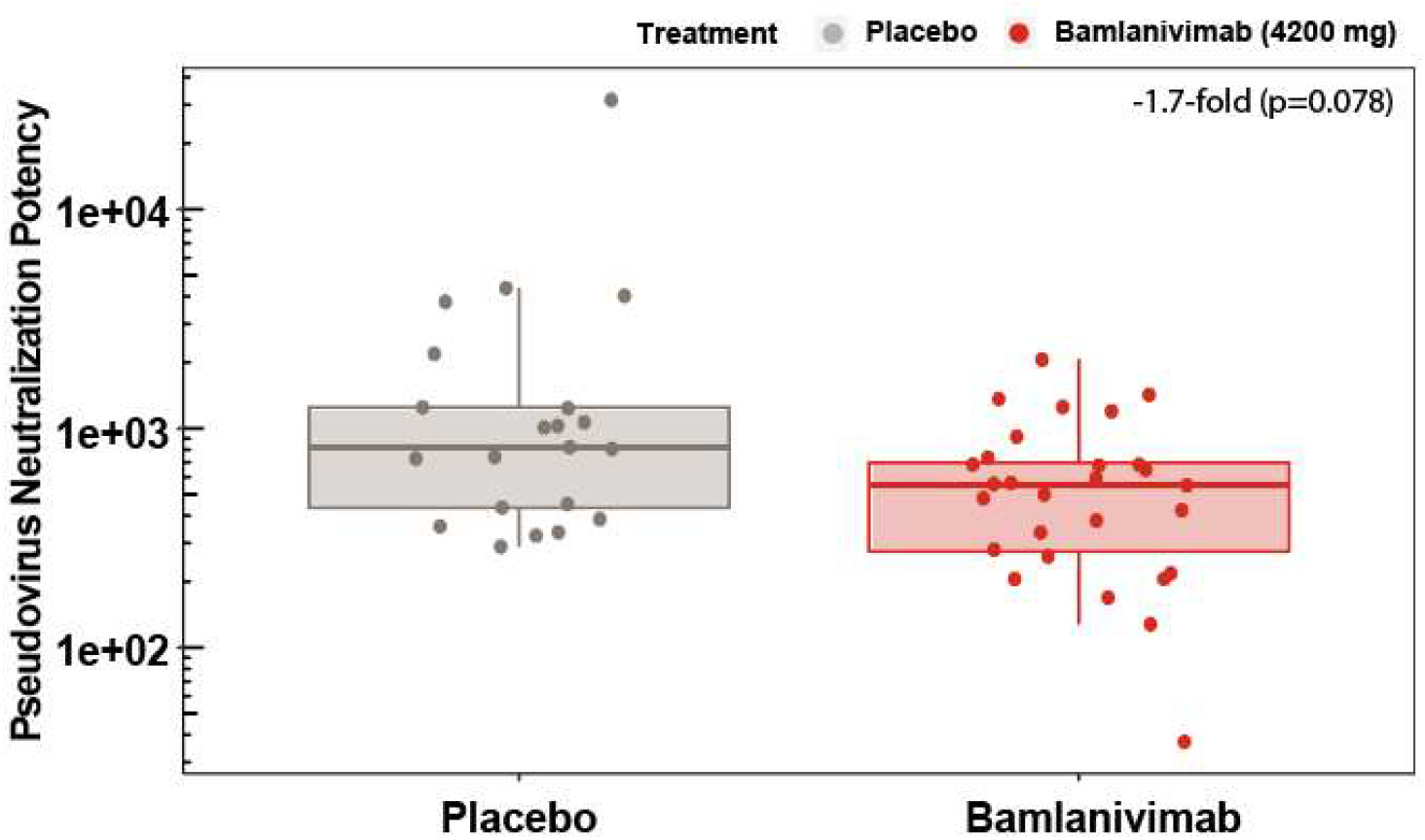
Pseudovirus neutralization potency (1/IC50) against Spike-E484Q. Samples collected from participants who received placebo or bamlanivimab and were subsequently fully vaccinated (N=49). Pseudovirus neutralization potency measured as 1/IC50 and adjusted for T1 and T2 covariates. Boxes and horizontal bars denote the interquartile range (IQR) and the median reciprocal IC50, respectively. Length of whiskers corresponds to 1.5 times the IQR.

We observed no difference in neutralization potency against Spike-E484Q between non-high-risk and high-risk staff (p=0.34), nor was the magnitude of the bamlanivimab effect on ACE2 binding inhibition potency different for these groups (p=0.085) (fig. S3b). To corroborate the results against Spike-E484Q pseudovirus, the neutralization potency was also evaluated against the beta variant (B.1.351) pseudovirus and similarly there was no statistically significant difference in the effect of bamlanivimab on antibody potency compared with placebo (−1.2-fold, p=0.465) (fig. S4a). Furthermore, there was a significantly strong Spearman correlation, ρ, of 0.8 (p<0.001) between the neutralization potency data against Spike-E484Q and the beta variant pseudoviruses (fig. S4b). Mirroring the findings with the other assays, there was no statistically significant difference in the effect of bamlanivimab on the pseudovirus neutralization potency against Spike-E484Q for participants, whether they were staff or resident (−2-fold, p=0.271) or whether they received Comirnaty or SpikeVax (−1.2-fold, p=0.819).

### Antibody titer, ACE2 binding inhibition and pseudovirus neutralization results show a high degree of correlation

To corroborate the results obtained using different assays to measure the antibody titers, ACE2 binding inhibition potency and pseudoviral neutralization potency, the correlation strength was determined between all assay results against Spike-RBD-E484Q for the subset of 114 samples from 49 participants. A high degree of correlation was observed between all assay results (Fig. 5). The Spearman correlation, ρ, was determined as 0.87 (p<0.001) for ACE2 binding inhibition data with pseudovirus neutralization data. Similarly, highly significant correlations were observed between pseudovirus neutralization data and antibody titers (ρ =0.84, p<0.001) and between ACE2 binding inhibition data and antibody titers (ρ =0.91, p<0.001). These strong correlations also extended to data from participants who received either bamlanivimab or placebo (Fig. 5).

**Fig. 5:**
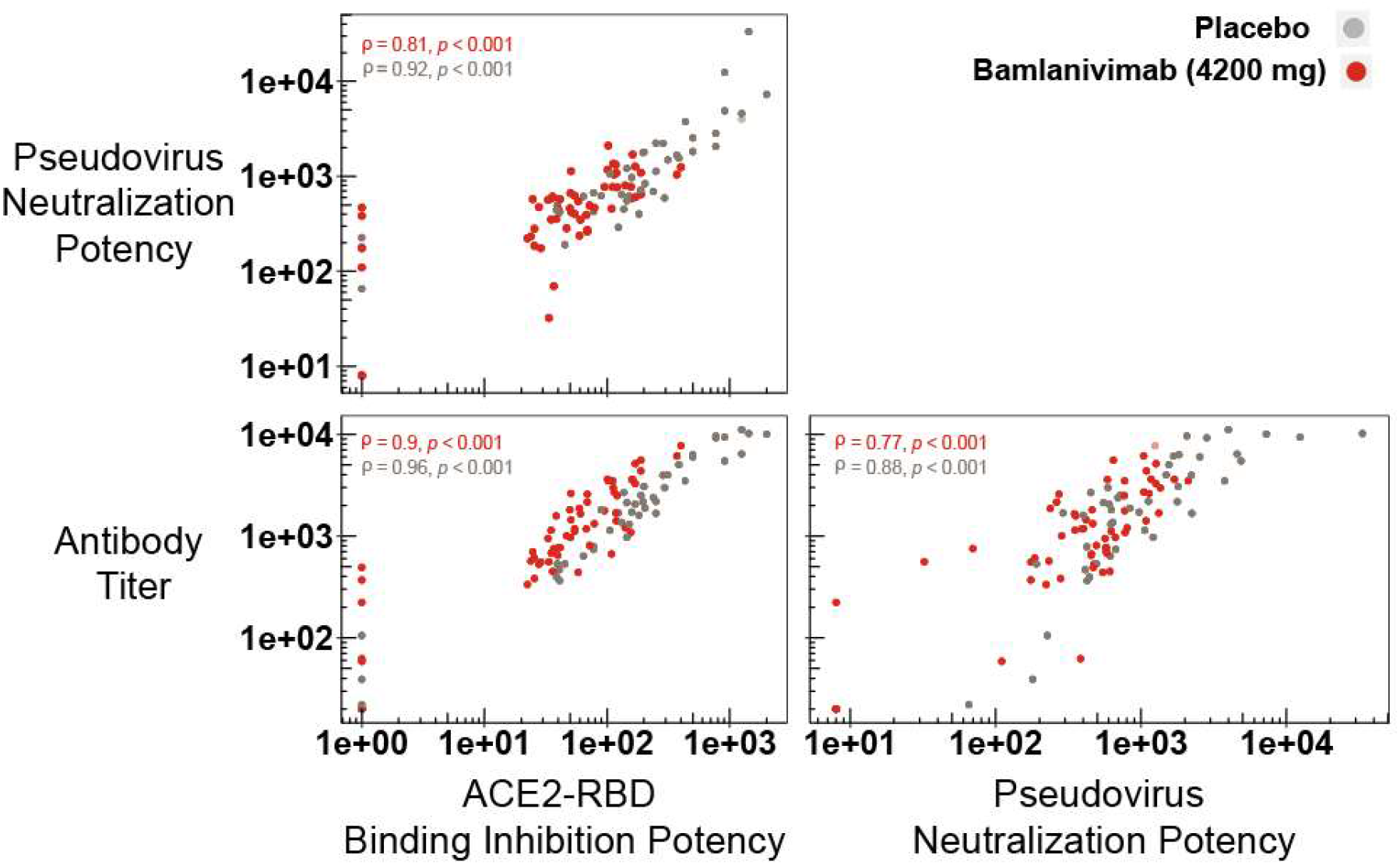
Correlation matrix showing the degree of correlation between paired results from assays: ACE2-RBD binding assay, pseudovirus assay (spike-E484Q) and antibody titers (spike-RBD-E484Q). Correlation strength determined for participants who received bamlanivimab or placebo. ρ represents the Spearman correlation; p represents the p-value.

### Longitudinal observation of antibody responses to COVID-19 vaccine

To visualize the longitudinal antibody responses to the COVID-19 vaccine, the antibody titers measured from all samples (n=499) from participants (N=135) were evaluated against Spike-RBD-E484Q. Since there was temporal variability in the number of days between receiving bamlanivimab or placebo and the first vaccine dose (from 43 - 127 days) the participants were divided into three groups based on the interval (T1) between bamlanivimab or placebo infusion and first vaccine dose. The longitudinal representation of the antibody titers against Spike-RBD-E484Q depicts the antibody response after the first COVID-19 vaccine dose for all fully vaccinated participants, whether infused with bamlanivimab or placebo (Fig. 6). The three groups were T1≤64 days, 64 days<T1≤85 days and T1>85 days with 50, 42 and 43 participants respectively (64 days and 85 days are the tertiles of T1). Samples collected within 2 weeks of the first vaccine dose from a subset of 27 participants (T1<64 days) showed that the median (range) bamlanivimab exposure at this time was 59.2 µg/mL (19.8-192.6 µg/mL). While the study did not allow for conclusions to be drawn about the immune response to the first vaccine dose, the data demonstrate that at no point in time did the groups divert beyond the small difference in titer observed at full vaccination. Additionally, there are no obvious differences between the immune responses shown in the three panels (Fig. 6). These trends were also observed for longitudinal antibody titers against the spike-NTD (fig. S5).

**Fig. 6:**
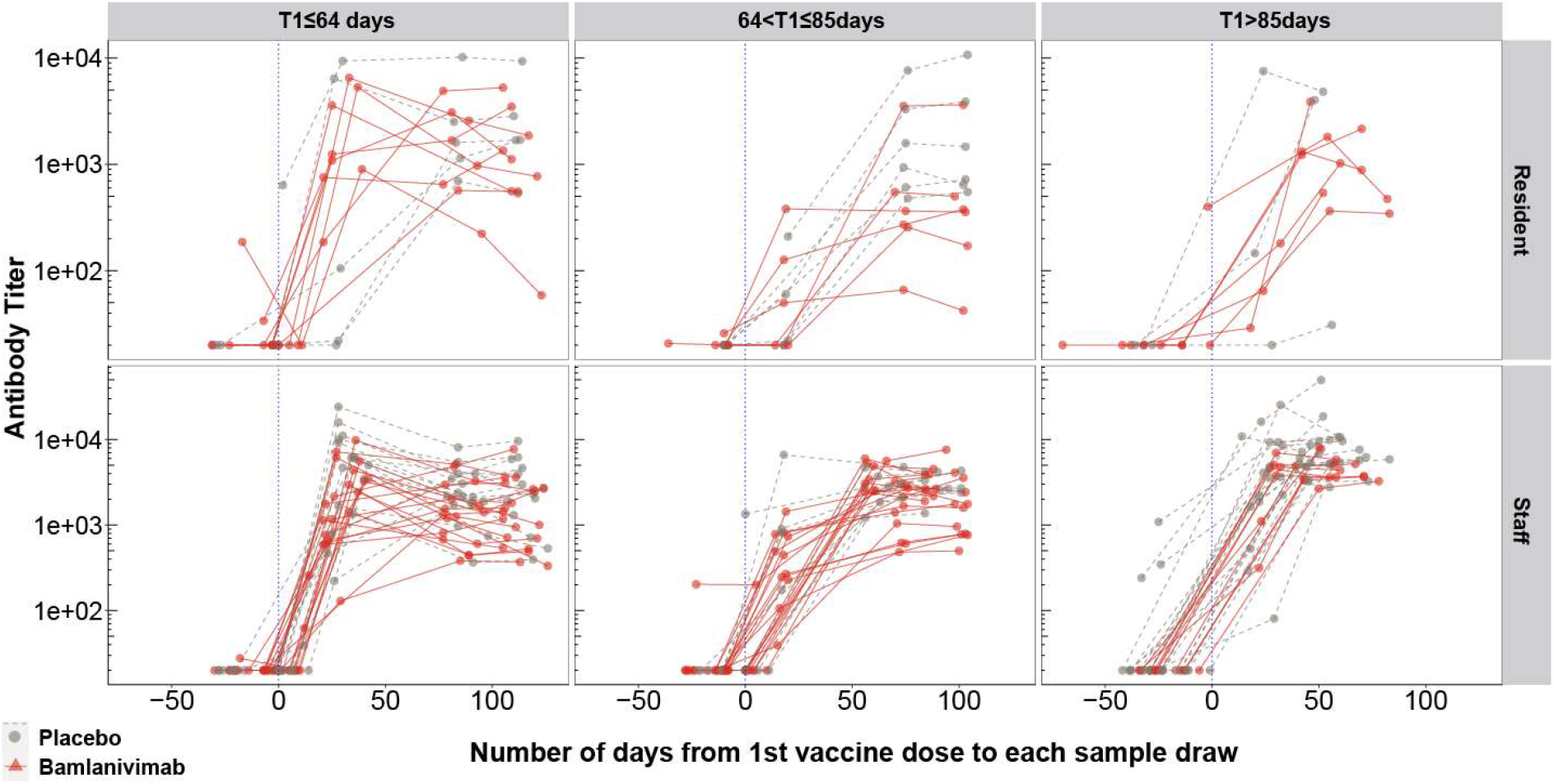
Longitudinal binding antibody responses against Spike-RBD-E484Q arranged into three groups based on tertiles of the interval (days) between bamlanivimab or placebo infusion and first vaccine dose, T1. Three columns (left to right) correspond to T1≤64 days, 64<T1≤85 days and T1>85 days, respectively. The vertical blue dotted line denotes the timepoint where participants receive the first dose of vaccine. Each line connects sample titers from a single participant. Top row shows antibody titers of participants who were residents and bottom row represents antibody titers of participants who were staff.

## Discussion

In this post-hoc analysis of the BLAZE-2 nursing home study, all participants demonstrated a robust immune response to full COVID-19 vaccination, regardless of preceding bamlanivimab or placebo infusion and irrespective of age, risk-category and vaccine type, with any observed differences unlikely to be clinically relevant. Furthermore, the interval between mAb infusion and COVID-19 vaccination did not affect this conclusion. These are significant findings in the pursuit of informed treatment planning, particularly for individuals at high risk for severe disease, and would support earlier COVID-19 vaccination for individuals who are currently deferring following mAb receipt, as per current CDC and WHO guidelines *(16, 17)*. Here, we demonstrate that SARS-CoV-2-naïve individuals who have received a prophylactic mAb can still mount a robust immune response to COVID-19 vaccination and therefore the benefit of prompt COVID-19 vaccination outweighs the minimal effect of a prior prophylactic COVID-19 mAb.

The immune response of participants to COVID-19 vaccination was evaluated using assays to measure the antibody titers, ACE2 binding inhibition and pseudoviral neutralization. There was a high degree of correlation between all assay results, suggesting reliable trends that demonstrated minimal differences in immune response to COVID-19 vaccination for participants who previously received either bamlanivimab or placebo. Notably, assays were measured against multiple domains of the spike protein that bamlanivimab does not significantly bind to, thereby reflecting the endogenous antibody response and with consistently strong correlations.

Growing evidence indicates that binding and neutralizing antibodies correlate with COVID-19 vaccine efficacy as many researchers endeavor to identify immune correlates of protection *(4, 18, 19)*. A recent modelling study identified a strong non-linear relationship between mean neutralization level and the reported protection of vaccines and predicted that the 50% protective neutralization level of a COVID-19 vaccine was achieved at approximately 20% of the mean convalescent titer *(31)*. Another study also estimated vaccine efficacy based on antibody marker level and showed that a 10-fold lower antibody titer for SpikeVax vaccine recipients, only reduced vaccine efficacy from 96.1% to 90.7% *(18)*. In our study antibody titers for fully vaccinated participants who had previously received bamlanivimab compared with placebo were reduced by 2-fold or less. Therefore, despite some differences reaching statistical significance, it is unlikely that a titer difference of this magnitude translates to clinically relevant interference. Higher antibody titers in individuals who were fully vaccinated with SpikeVax compared with Comirnaty have been reported *(32)*, yet each vaccine has demonstrated more than 90% efficacy in preventing COVID-19 illness *(33, 34)*. We concluded here that there was no significant difference in the antibody response to different mRNA COVID-19 vaccines and that bamlanivimab infusion also did not disparately affect participants who had either vaccine.

Understanding the endogenous antibody responses to COVID-19 vaccines is particularly important for older and immunocompromised individuals, who are at high-risk of developing severe COVID-19 and have therefore been the targeted recipients of prophylactic and treatment mAbs *(35-38)*. The participants in this study included both residents and staff of US skilled nursing and assisted living facilities, allowing us to evaluate any impact of age and risk categorization on the vaccine-induced antibody response following receipt of prophylactic bamlanivimab or placebo. Firstly, staff (median age 43 years) were shown to have significantly higher antibody titers and greater antibody potency than residents (median age 72 years), which is consistent with the literature that has shown stronger immune responses in younger individuals *(39)*. Secondly, bamlanivimab had a similar effect on the antibody titers and potency in both residents and staff, which informs that bamlanivimab did not disparately affect the immune response to full COVID-19 vaccination of participants of different ages. An additional sub-analysis was completed for the staff who were dichotomized into non-high-risk and high-risk participants based on pre-determined risk factors for developing severe COVID-19 disease *(13)*. Since only 13% of the high-risk staff were 65 years or older, the comparative differences between these risk groups were largely attributable to factors other than older age. Firstly, no differences in antibody titer or potency were observed between the two risk-groups, and bamlanivimab infusion also did not disparately affect the titers and potency of elicited antibodies in participants of different risk categorization. These results therefore demonstrate that participants can mount a strong immune response to COVID-19 vaccination following a mAb infusion, irrespective of age and high-risk categorization. This is an important finding as mAbs have been granted EUAs primarily for the treatment of individuals who are at high risk of developing severe COVID-19 and the current lack of data resulted in substantial delays between COVID-19 mAb and vaccination for this group. A further consideration based on the reduced protective efficacy of COVID-19 vaccines over time and against certain SARS-CoV-2 variants, additional doses are increasingly administered to boost the immune response, with high-risk individuals being prioritized *(40)*. Taken together, these data reinforce the conclusions, that individuals who have previously received a COVID-19 mAb can proceed with COVID-19 vaccination and mount a strong immune response, which could be further boosted by additional vaccine doses, as per the latest clinical guidance.

The vaccine-induced antibody potency was also evaluated using an ACE2 binding inhibition assay and a pseudovirus neutralization assay with closely correlated results. There was no significant difference in pseudovirus neutralization potency between individuals who received either bamlanivimab or placebo, however a significantly lower ACE2 binding inhibition potency was detected. The contrast in measured antibody potency is likely explained by the breadth of epitope assessed in each assay; the ACE2 binding inhibition assay assesses RBD-binding antibodies only, whereas the pseudovirus assay assesses the functionality of the polyclonal antibody response against the full-length spike (see minimum significant ratio in Materials and Methods for full description). The strong correlation between the potency assays and overall high levels of neutralizing activity suggests minimal impact on immune protection conferred by COVID-19 vaccination, regardless of prior bamlanivimab or placebo infusion. Previous studies investigating immune evasion have determined that complete loss of antibody neutralizing activity against SARS-CoV-2 variants corresponded to a greater than 40-fold change reduction compared with neutralizing activity against wild type (WT) pseudovirus *(41, 42)*. Therefore, the decrement in antibody titers observed in participants who received prior bamlanivimab infusion compared with placebo is unlikely to be clinically meaningful.

This study has several limitations; First, this analysis was not a pre-planned component of the BLAZE-2 trial and therefore vaccine type and timing were determined by circumstance. Consequently, the post-hoc analysis population was determined as described in the Materials and Methods and therefore the data presented are limited by the sample size and demographics. A total of 499 samples from fully vaccinated participants who met the inclusion criteria were assessed for antibody titer and ACE2 binding inhibition potency using a custom Luminex-based assay. Owing to the custom nature of this assay, it was decided to perform a standard pseudovirus neutralization assay to complement and corroborate these data. Due to logistical limitations, purposive sampling was used to select samples for the pseudovirus assay from a subset of participants (N=49) who received their first vaccine within 64 days of either bamlanivimab or placebo. This group of participants were selected as they represented those most likely to exhibit an effect of bamlanivimab on pseudovirus neutralization potency. Despite the smaller sample, the neutralization potency against Spike-RBD-E484Q and the beta variant pseudoviruses were strongly correlated, further supporting our interpretation of minimal impact.

This analysis only assessed the impact of a single mAb on the endogenous immune response to a COVID-19 vaccine, however, we hypothesize similar results for other mAbs that reduce viral load upon administration *(43)*. This study is also limited to participants who received an mRNA vaccine; therefore, it is not known whether these findings extend to other vaccine types. This analysis also did not assess the simultaneous administration of vaccine and mAb, however we have recently shown that patients with mild or moderate COVID-19 that were administered bamlanivimab or bamlanivimab and etesevimab together early in infection elicited a wide breadth of antigenic responses to SARS-CoV-2 *(44)*.

Since COVID-19 vaccination was an unplanned component of the BLAZE-2 trial, the temporal variability of vaccination dosing relative to the serum sampling visits had to be accounted for throughout this post-hoc analysis. The most appropriate way to compare the immune responses of participants to vaccination was to adjust the data of each participant for two covariates (Materials and Methods). Longitudinal titer data without covariate adjustment were also visualized by arranging participants into three groups based on the interval between bamlanivimab or placebo infusion and first vaccine dose. Although this study did not allow for conclusions to be drawn about the immune response to the first dose of vaccine, the longitudinal titer data did not deviate beyond the small difference in titer observed at full vaccination. Whilst this approach was limited by the number of participants who could be included in each group, there were no obvious differences in the antibody responses between the three groups, which notably included 97 participants (70%) who had received the first vaccine dose within 90 days of bamlanivimab or placebo infusion *(16, 17)*.

In conclusion, this post-hoc analysis expands the current understanding of the impact of receiving a prophylactic monoclonal antibody infusion, along with other factors, on the endogenous immune response to full COVID-19 vaccination. There was a high degree of correlation between all assay results of vaccine-induced antibody titer and potency against different SARS-CoV-2 proteins supporting the conclusion that participants mount a strong immune response to full COVID-19 vaccination, regardless of preceding prophylactic mAb infusion and irrespective of age, risk-category and vaccine type. The observed incremental differences are unlikely to be clinically consequential. These findings are pertinent for informing public health policy, particularly for SARS-CoV-2-naïve individuals and those at high-risk of developing severe COVID-19 illness, who can receive immediate protective immunity from COVID-19 mAbs while awaiting the development of durable polyclonal vaccine-induced protection. Results also demonstrate that the benefit of receiving a COVID-19 vaccination at the earliest opportunity outweighs any minimal effect on the endogenous immune response due to prior COVID-19 mAb infusion. These data therefore advance the understanding of the current COVID-19 treatment and prevention portfolio and indicate that COVID-19 mAbs can play a complementary, rather than a competing role with COVID-19 vaccines.

## Materials and Methods

### Experimental Design

The BLAZE-2 trial was a phase 3, randomized, double-blind, placebo-controlled, single-dose SARS-CoV-2 prevention study and has been described previously *(22)*. Participants of the BLAZE-2 study included residents and staff of US skilled nursing and assisted living facilities who were randomized to the study drug, bamlanivimab (4,200 mg) or placebo. As per the trial protocol, serum samples were collected from participants at baseline (prior to bamlanivimab or placebo infusion) and post-baseline samples were collected at day 29, day 57, day 85, day 141 and day 169 *(22)*. All donors provided written informed consent for the use of blood and blood components (such as PBMCs, sera or plasma). In an unscheduled component of this study, a subset of participants also received two doses of a mRNA COVID-19 vaccine (Comirnaty or SpikeVax) as part of the US vaccination program during this sampling period. At the time of receipt, the Comirnaty and SpikeVax mRNA vaccines had been granted Emergency Use Authorizations (EUAs) by the US Food and Drug Administration (FDA) *(34, 45)*.

The selection process for participant inclusion in this post-hoc analysis is presented in Fig. 7. The participant sample size for this post-hoc analysis was therefore dictated by circumstance and included BLAZE-2 participants who met all of the following criteria: (a) participants were in the prevention cohort of the BLAZE-2 trial; (b) tested negative for SARS-CoV-2 throughout the study, as determined using both reverse transcription polymerase chain reaction (RT-PCR; assessed at baseline and then weekly until day 57 and on days 85 and 141) and nucleocapsid (NCP) antibody assay (Cobas, Roche Diagnostics; assessed on days 1, 29, 57, 85, and 141) (c) and had received two COVID-19 vaccine doses subsequent to a bamlanivimab or placebo infusion and (d) had at least one serum sample obtained more than 2 weeks following the second vaccine dose. The CDC describes an individual as fully vaccinated after 2 weeks following the second COVID-19 vaccine doses in a 2-dose series, such as for Comirnaty or SpikeVax *(23)*. The baseline characteristics for the 135 participants who met these criteria and were included in this post-hoc analysis are presented in Table 1.

**Fig. 7:**
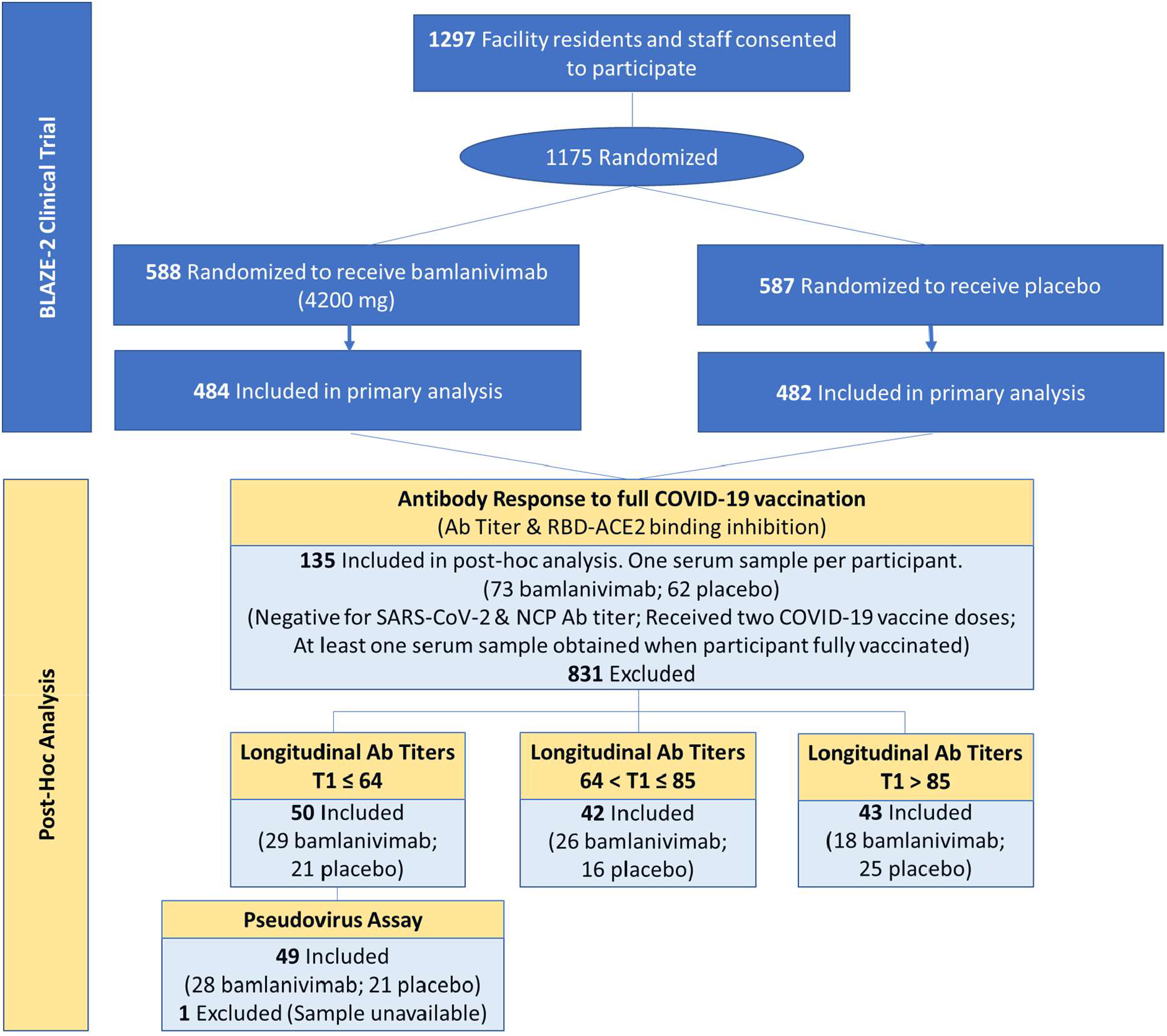
Selection of fully vaccinated participants for post-hoc analysis. *COVID-19 Vaccines: Comirnaty (Pfizer/BioNTech); SpikeVax (Moderna). T1 = interval (days) between bamlanivimab or placebo infusion and first COVID-19 vaccine dose. The CDC describes an individual as fully vaccinated after 2 weeks following the second COVID-19 vaccine dose in a 2-dose series, such as for Comirnaty or SpikeVax (23)*.

Participants received COVID-19 vaccines (SpikeVax or Comirnaty) when they were offered to the respective nursing and assisted living facilities by the US government. Consequently, participants received a first vaccine dose at different timepoints (starting at day 44 onwards) following the bamlanivimab or placebo infusion. Participants received the second vaccine dose following the recommended period specified in the EUA factsheet for each vaccine (21 and 28 days later for Comirnaty and SpikeVax, respectively) *(25, 26)*.

To evaluate the effect of bamlanivimab infusion on the subsequent antibody response to full COVID-19 vaccination, the following criteria were adopted to select samples from the 135 participants for statistical analysis: (a) exclude participant samples that were obtained prior to the receipt of the second vaccine dose; (b) exclude samples from participants who did not have a record of a second vaccine dose at the time of the analysis; (c) exclude samples which were obtained within 14 days of the second vaccine dose (i.e. only use samples collected after full vaccination, as determined by the CDC *(23)*; (d) If more than one sample was obtained after a participant was fully vaccinated, the lattermost sample was selected for analysis. The serum sampling period was pre-specified in the BLAZE-2 protocol and the final serum samples were collected on day 169 *(22)*. A total of 499 samples from 135 participants met these criteria and assays were performed to measure antibody titers and ACE2-RBD binding inhibition as described in subsequent sections. Prior to use in each assay, serum samples were centrifuged for 5 minutes at 10000 x *g* to pellet any debris.

Since the ACE2-RBD binding inhibition results were collected using a custom Luminex-based assay, a standard VSV pseudoviral assay was also performed to complement and corroborate these data. Owing to the substantial number of serum samples collected, the pseudoviral assay was performed on a purposive sample of 74 samples from 49 fully vaccinated participants (Fig. 7).

Longitudinal analysis of antibody titers measured from all 499 samples obtained from the 135 participants facilitated the visualization of the antibody response by each individual to COVID-19 vaccination. Since the timing of vaccine dosing varied for each individual, the participants were organized into three groups based on the interval between receipt of bamlanivimab or placebo and the subsequent receipt of the first COVID-19 vaccination dose, T1. Each of the 135 participants were placed into one of the three groups: T1≤64 days, 64<T1<85 days and T1≥85 days.

### Custom Luminex-Based Assay

Luminex xMAP technology is an established, multiplex, flow cytometry-based platform that allows the simultaneous quantitation of many protein analytes in a single reaction *(46)*. A custom Luminex-based assay was developed to measure serology and antibody ACE2-RBD binding inhibition in a single assay. Antigen-coated microspheres were used to detect and quantitate endogenous antibodies against SARS-CoV-2 proteins, including spike-NTD and several RBD epitopes (Table 2), to which bamlanivimab does not significantly bind *(7, 47)*.

Patient serum samples were titrated (1:20 – 1: 4.3E8) in phosphate buffered saline-high salt solution (PBS-HS; 0.01 M PBS, 1% [bovine serum albumin] BSA, 0.02% Tween, 300 mM NaCl). Diluted serum samples were combined with Luminex MAGPlex microspheres coupled with individual antigens and a recombinant, labelled RBD-PE protein and incubated for 60 minutes to allow endogenous antibodies to bind to either the recombinant RBD (E484Q)-PE or to the antigen-coated Luminex beads. The solution was placed on a magnet, collecting the MAGPlex beads, while the supernatant was transferred to a new plate. The transferred solution was combined with ACE2 coated beads and incubated for 60 minutes, while the remaining beads were washed and incubated for 60 minutes with anti-IgG-PE beads to detect bound antibodies.

All the beads on both plates were then washed and resuspended in a PBS-1% BSA solution and read using a Luminex FlexMAP 3D System with xPONENT Software. The titer was evaluated from the median fluorescence intensity (MFI) and the ability of the endogenous antibodies to inhibit RBD (E484Q)-ACE2 binding was calculated based on the half maximal inhibitory concentration, IC50, which represents the antibody titer where the ACE2-RBD (E484Q) binding is reduced by half. The ACE2 binding inhibition potency was assessed using the inverse of IC50.

### Pseudovirus production and characterization

E484Q mutagenesis reactions were performed using the QuickChange Lightning Site-Directed Mutagenesis Kit (Agilent #210519) using a template of a spike mammalian expression vector based on the Wuhan sequence (Genbank MN908947.3) with a deletion of the C-terminal 19 amino acids. For the Beta variant (B.1.351) pseudovirus a consensus sequence representative of lineage was synthesized and incorporated by Gibson cloning. Pseudoviruses bearing mutant spike proteins were produced using the delta-G-luciferase recombinant Vesicular Stomatitis Virus (rVSV) system (KeraFast EH1025-PM, Whitt 2010). Briefly, 293T cells were transfected with individual mutant spike expression plasmids, and 16-20 hours later, transfected cells were infected with VSV-G-pseudotyped delta-G luciferase rVSV, and 16-20 hours thereafter conditioned culture medium was harvested, clarified by centrifugation at 1320 g for 10 minutes at 4°C, aliquoted and stored frozen at - 80°C. Relative luciferase reporter signal read-out was determined by luciferase assay (Promega E2650) of extracts from VeroE6 cells infected with serially-diluted virus. Luciferase activity was measured on a PerkinElmer EnVision 2104 Multilabel Reader.

### Pseudovirus neutralization assays

Neutralization assays were carried out essentially as described previously *(48, 49)*. Serum antibodies were diluted 4-fold in assay media and 10-point 3-fold titrations in 25% assay media were performed in 384-well polystyrene plates in duplicate using a Beckman (Biomek i5) liquid handler. Positive and negative control antibodies and an unrelated control (hIgG1 isotype) were tested in a 10-point, 3-fold serial dilution starting at 8 µg/mL, 2 µg/mL and 8 µg/mL, respectively, in 25% assay media. An empirically pre-determined fixed amount of pseudovirus (Spike-RBD-E484Q or the beta variant (B.1.351) spike; Table 2) was dispensed by WDII liquid dispenser on titrated serum antibodies and controls and pre-incubated for 20 minutes at 37°C. Following pre-incubation, the virus-antibody complexes were transferred by Biomek i5 to 8,000/well VeroE6 cells in white, opaque, tissue culture treated 384W plates, and incubated for 16-20 hours at 37°C. Control wells included virus only (no antibody; 14 replicates) and cells only (14 replicates). Following infection, cells were lysed with Promega BrightGlo and luciferase activity was measured on the Biotek Synergy Neo2 Multimode Reader. Antibody potency was assessed using the inverse of IC50, defined as the antibody concentration, at which the viral replication has been reduced by 50% relative to the absence of antibodies.

### Statistical Analysis

For the serial dilution-based serology assay, titer can either be defined as the reciprocal of the highest dilution of the sample above a pre-determined “cut point” value or be derived based on interpolating assay values that straddle the “cut point” *(50)*. The latter method was used in the forementioned serology assay. The serology titer data were evaluated on a log base 10 scale.

To calculate IC50 of data from the pseudovirus neutralization assay and from the ACE2 neutralization component of the serology assay, a 4-parameter logistic function was used to estimate the absolute IC50 based on 1/dilution factor (bottom is fixed at 0 for pseudovirus IC50). For the pseudovirus neutralization assay, if a sample indicates no neutralization or has a poor fit (the standard error of the IC50 is not estimable or the estimated IC50 is larger than the maximum 1/dilution factor), the IC50 value was imputed as 0.125 (twice the maximum 1/dilution factor). For the ACE2 neutralization assay, if a sample indicates no neutralization, the IC50 was imputed to 1 (20 times the maximum 1/dilution factor). For both neutralization assays analyses, bamlanivimab effect (compared with placebo) was evaluated based on log_10_(1/IC50).

Since COVID-19 vaccination was not a planned component of this study, the temporal variability in vaccine dosing for each participant relative to the sampling schedule had to be accounted for in the analysis. For each participant, let T1 denote the interval (days) between bamlanivimab or placebo infusion and first COVID-19 vaccine dose and T2 denote the interval (days) from second COVID-19 vaccine dose to each sampling visit. T1 and T2 were included as covariates in the linear model used for hypothesis testing (2-sided test with α level of 0.05). To compare results from participants who received either bamlanivimab or placebo infusion, the respective treatment group was included in the linear model. To compare the effect of bamlanivimab or placebo infusion on the results from residents or staff, treatment group (bamlanivimab or placebo), patient group (resident or staff) and treatment x patient group interaction were included in the linear model. To compare results from participants who received different vaccines, the treatment group (bamlanivimab or placebo), vaccine type (Comirnaty or SpikeVax), and treatment x vaccine type interaction were included in the linear model. Adjustments for multiple testing were not conducted; therefore, the findings should be interpreted as exploratory. The statistical analyses were performed with R software (version 4.0.3) *(51)*.

To visualize the results after adjusting for T1 and T2, residuals adjusted for T1 and T2 (Equation 1) were rescaled to either titer or 1/IC50 scales, depending on which response variable was evaluated in the linear model, where Y = either log_10_(titer) or log_10_(1/IC50), Let Y^*^ denote the rescaled Y. Since Y ∼ N(µY, σY ^2^ and 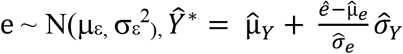, where the “*hat*” indicates the estimated value.

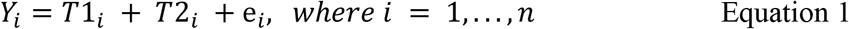

### Minimum Significant Ratio of Assay Variability

The minimum significant ratio (MSR) is a statistical parameter used to measure assay variability *(52)*. The replicate experiment MSR was determined as described in *(53)* for the ACE2 binding inhibition assay and pseudovirus assay used in this post-hoc analysis. Pseudovirus assay exhibits higher variability (MSR ≈ 5) compared to ACE2-RBD assay (MSR ≈ 1.2), which reflects the breadth of epitope assessed in each assay. The ACE2 binding inhibition assay has greater precision as it assesses RBD-binding antibodies only, whereas the pseudovirus assay assesses the functionality of the polyclonal antibody response against the full-length spike.

## Data Availability

All data produced in the present work are contained in the manuscript

## Supplementary Materials

**Figure S1:** Antibody titers against SARS-CoV-2 beta variant (B.1.351) for fully vaccinated participants who (a) previously received either bamlanivimab or placebo infusion (b) who were resident of staff and were subsequently fully vaccinated (SpikeVax or Comirnaty) against COVID-19.

**Figure S2:** Antibody titers against (a) Spike-RBD-E484Q and (b) spike-NTD comparing samples from fully vaccinated participants who were non-high-risk staff and high-risk staff (top) and further grouped by those who received placebo or bamlanivimab prior to vaccination (bottom).

**Figure S3:** Antibody potency (1/IC50) as measured using (a) ACE2 binding inhibition potency and (b) Spike-RBD-E484Q pseudovirus neutralization for fully vaccinated staff participants who were categorized as non-high risk or high risk (top) and further grouped by those who received placebo or bamlanivimab prior to vaccination.

**Figure S4:** (a) Neutralization potency (1/IC50) against SARS-CoV-2 beta variant (B.1.351) pseudovirus measured for samples collected from participants (N=49) who received placebo or bamlanivimab and were subsequently fully vaccinated. (b) Correlation plot of neutralization potency against beta variant and against E484Q

**Figure S5:** Longitudinal antibody responses against spike-NTD arranged into three groups based on the interval (days) between bamlanivimab or placebo infusion and first vaccine dose, T1.

## Trial Registration

ClinicalTrials.gov Identifier: NCT04497987 (Note: The results herein were not a pre-planned component of this trial).

## Acknowledgements

We would like to thank John Mascola, MD, for his valuable advice and assessment of the data. We would also like to thank Inmaculada de la Torre for her review and comments. We thank AbCellera Biologics for their collaboration in developing bamlanivimab. Bamlanivimab emerged from the collaboration between Eli Lilly and Co and AbCellera Biologics to create antibody therapies for the prevention and treatment of COVID-19. Eli Lilly and Co developed the antibody after it was discovered by AbCellera and scientists at the NIAID Vaccine Research Center.) We also thank the National Institutes of Health, the study participants, and the support staff, whose contributions were vital to this project.

## Author contributions

Conceptualization: RJB, JLT, REH, LZ.

Study Design: RJB, JLT, LZ, JP, MD, SB, TNVT, MC, JJF, PV, NLK, REH and AN.

Data Acquisition: JLT, JP, PV, MD, SB.

Data Interpretation: RJB, JLT, JP, LZ, REH, MC, NLK, NH, MSC and MM.

Data Analysis: LZ, MC, TNVT, REH.

Writing: RJB, JLT, JP, LZ, NH, MSC and MM.

All authors revised the manuscript.

All authors read and approved the final version for submission.

## Conflict of Interest

All authors have completed the ICMJE uniform disclosure form at www.icmje.org/coi_disclosure.pdf and declare: This work was supported by Eli Lilly and Company. RJB, JLT, LZ, NLK, JP, SB, TNVT, MC, JJF, MD PV, LZ, REH and AN are all employees and stockholders of Eli Lilly and Company. NH is an employee of Eli Lilly and Company. MSC declared study funding by National Institute of Health (NIH), consulting fees from Lineage Logistics and Atea Pharmaceutical, payment for educational events from NY Course, Magen, MJH Life Sciences, Clinical Care solutions and Virology Education, support for attending advisory meetings from Krog Healthcare – GlaxoSmithKline (GSK), Trinity Health Sciences and GSK, leadership or fiduciary roles at HPTN, CoVPN, Fogarty and McGill.

## Notes

### Clinical Protocols

https://jamanetwork.com/journals/jama/fullarticle/2780870

### Funding Statement

This study was funded by Eli Lilly and Company.

### Author Declarations

IRB of Advarra gave ethical approval for the entire BLAZE-2 study on July 2020 and all institutional review boards also provided ethical approval.

## References

1. Hu, B., H. Guo, P. Zhou, and Z.-L. Shi, Characteristics of SARS-CoV-2 and COVID-19. Nature Reviews Microbiology, 2021. 19(3): p. 141–154.

2. Siemieniuk, R.A., J.J. Bartoszko, L. Ge, D. Zeraatkar, A. Izcovich, E. Kum, H. Pardo-Hernandez, A. Qasim, J.P.D. Martinez, B. Rochwerg, F. Lamontagne, M.A. Han, Q. Liu, A. Agarwal, T. Agoritsas, D.K. Chu, R. Couban, E. Cusano, A. Darzi, T. Devji, B. Fang, C. Fang, S.A. Flottorp, F. Foroutan, M. Ghadimi, D. Heels-Ansdell, K. Honarmand, L. Hou, X. Hou, Q. Ibrahim, A. Khamis, B. Lam, M. Loeb, M. Marcucci, S.L. McLeod, S. Motaghi, S. Murthy, R.A. Mustafa, J.D. Neary, G. Rada, I.B. Riaz, B. Sadeghirad, N. Sekercioglu, L. Sheng, A. Sreekanta, C. Switzer, B. Tendal, L. Thabane, G. Tomlinson, T. Turner, P.O. Vandvik, R.W. Vernooij, A. Viteri-García, Y. Wang, L. Yao, Z. Ye, G.H. Guyatt, and R. Brignardello-Petersen, Drug treatments for covid-19: living systematic review and network meta-analysis. BMJ, 2020. 370: p. m2980.

3. Robbiani, D.F., C. Gaebler, F. Muecksch, J.C.C. Lorenzi, Z. Wang, A. Cho, M. Agudelo, C.O. Barnes, A. Gazumyan, S. Finkin, T. Hägglöf, T.Y. Oliveira, C. Viant, A. Hurley, H.-H. Hoffmann, K.G. Millard, R.G. Kost, M. Cipolla, K. Gordon, F. Bianchini, S.T. Chen, V. Ramos, R. Patel, J. Dizon, I. Shimeliovich, P. Mendoza, H. Hartweger, L. Nogueira, M. Pack, J. Horowitz, F. Schmidt, Y. Weisblum, E. Michailidis, A.W. Ashbrook, E. Waltari, J.E. Pak, K.E. Huey-Tubman, N. Koranda, P.R. Hoffman, A.P. West, C.M. Rice, T. Hatziioannou, P.J. Bjorkman, P.D. Bieniasz, M. Caskey, and M.C. Nussenzweig, Convergent antibody responses to SARS-CoV-2 in convalescent individuals. Nature, 2020. 584(7821): p. 437–442.

4. Earle, K.A., D.M. Ambrosino, A. Fiore-Gartland, D. Goldblatt, P.B. Gilbert, G.R. Siber, P. Dull, and S.A. Plotkin, Evidence for antibody as a protective correlate for COVID-19 vaccines. Vaccine, 2021. 39(32): p. 4423–4428.

5. Legros, V., S. Denolly, M. Vogrig, B. Boson, E. Siret, J. Rigaill, S. Pillet, F. Grattard, S. Gonzalo, P. Verhoeven, O. Allatif, P. Berthelot, C. Pélissier, G. Thiery, E. Botelho-Nevers, G. Millet, J. Morel, S. Paul, T. Walzer, F.-L. Cosset, T. Bourlet, and B. Pozzetto, A longitudinal study of SARS-CoV-2-infected patients reveals a high correlation between neutralizing antibodies and COVID-19 severity. Cellular & Molecular Immunology, 2021. 18(2): p. 318–327.

6. Pujadas, E., F. Chaudhry, R. McBride, F. Richter, S. Zhao, A. Wajnberg, G. Nadkarni, B.S. Glicksberg, J. Houldsworth, and C. Cordon-Cardo, SARS-CoV-2 viral load predicts COVID-19 mortality. The Lancet. Respiratory medicine, 2020. 8(9): p. e70–e70.

7. Gottlieb, R.L., A. Nirula, P. Chen, J. Boscia, B. Heller, J. Morris, G. Huhn, J. Cardona, B. Mocherla, V. Stosor, I. Shawa, P. Kumar, A.C. Adams, J. Van Naarden, K.L. Custer, M. Durante, G. Oakley, A.E. Schade, T.R. Holzer, P.J. Ebert, R.E. Higgs, N.L. Kallewaard, J. Sabo, D.R. Patel, P. Klekotka, L. Shen, and D.M. Skovronsky, Effect of Bamlanivimab as Monotherapy or in Combination With Etesevimab on Viral Load in Patients With Mild to Moderate COVID-19: A Randomized Clinical Trial. JAMA, 2021.

8. Slifka, M.K. and I.J. Amanna, 8 - Passive Immunization, in Plotkin’s Vaccines (Seventh Edition), S.A. Plotkin, et al., Editors. 2018, Elsevier. p. 84-95.e10.

9. US Food and Drug Administration, Fact sheet for health care providers: Emergency Use Authorization (EUA) of bamlanivimab and etesevimab 2021, Accessed 30 September 2021: https://www.fda.gov/media/145802/download.

10. US Food and Drug Administration, Fact sheet for health care providers: Emergency Use Authorization (EUA) of caririvimab and imdevimab. 2021: https://www.fda.gov/media/143892/download.

11. US Food and Drug Administration, Fact sheet for health care providers: Emergency Use Authorization (EUA) of bamlanivimab 2020: https://www.fda.gov/media/143603/download

12. US Food and Drug Administration, Coronavirus (COVID-19) Update: FDA Authorizes Additional Monoclonal Antibody for Treatment of COVID-19. 2021. May, 26: https://www.fda.gov/news-events/press-announcements/coronavirus-covid-19-update-fda-authorizes-additional-monoclonal-antibody-treatment-covid-19.

13. Chen, P., A. Nirula, B. Heller, R.L. Gottlieb, J. Boscia, J. Morris, G. Huhn, J. Cardona, B. Mocherla, V. Stosor, I. Shawa, A.C. Adams, J. Van Naarden, K.L. Custer, L. Shen, M. Durante, G. Oakley, A.E. Schade, J. Sabo, D.R. Patel, P. Klekotka, and D.M. Skovronsky, SARS-CoV-2 Neutralizing Antibody LY-CoV555 in Outpatients with Covid-19. New England Journal of Medicine, 2020. 384(3): p. 229–237.

14. Clem, A.S., Fundamentals of vaccine immunology. Journal of global infectious diseases, 2011. 3(1): p. 73–78.

15. Sparrow, E., M. Friede, M. Sheikh, and S. Torvaldsen, Therapeutic antibodies for infectious diseases. Bulletin of the World Health Organization, 2017. 95(3): p. 235–237.

16. Centers for Disease Control and Prevention, Interim Clinical Considerations for Use of COVID-19 Vaccines Currently Authorized in the United States. 2021, Mar; [Accessed 2021, Nov 9]: https://www.cdc.gov/vaccines/covid-19/info-by-product/clinical-considerations.html.

17. World Health Organization, Interim recommendations for use of the Pfizer–BioNTech COVID-19 vaccine, BNT162b2, under Emergency Use Listing. 8 January, 2021: https://apps.who.int/iris/bitstream/handle/10665/338484/WHO-2019-nCoV-vaccines-SAGE_recommendation-BNT162b2-2021.1-eng.pdf [Accessed Nov 9, 2021].

18. Gilbert, P.B., D.C. Montefiori, A. McDermott, Y. Fong, D. Benkeser, W. Deng, H. Zhou, C.R. Houchens, K. Martins, L. Jayashankar, F. Castellino, B. Flach, B.C. Lin, S. O’Connell, C. McDanal, A. Eaton, M. Sarzotti-Kelsoe, Y. Lu, C. Yu, B. Borate, L.W.P. van der Laan, N. Hejazi, C. Huynh, J. Miller, H.M. El Sahly, L.R. Baden, M. Baron, L. De La Cruz, C. Gay, S. Kalams, C.F. Kelley, M. Kutner, M.P. Andrasik, J.G. Kublin, L. Corey, K.M. Neuzil, L.N. Carpp, R. Pajon, D. Follmann, R.O. Donis, R.A. Koup A. on behalf of the Immune, I. Moderna, E. Coronavirus Vaccine Prevention Network /Coronavirus, and V.P.N.B.T. United States Government /Co, Immune Correlates Analysis of the mRNA-1273 COVID-19 Vaccine Efficacy Trial. medRxiv, 2021: p. 2021.08.09.21261290.

19. Corbett Kizzmekia, S., C. Nason Martha, B. Flach, M. Gagne, S. O’Connell, S. Johnston Timothy, N. Shah Shruti, V. Edara Venkata, K. Floyd, L. Lai, C. McDanal, R. Francica Joseph, B. Flynn, K. Wu, A. Choi, M. Koch, M. Abiona Olubukola, P. Werner Anne, I. Moliva Juan, F. Andrew Shayne, M. Donaldson Mitzi, J. Fintzi, R. Flebbe Dillon, E. Lamb, T. Noe Amy, T. Nurmukhambetova Saule, J. Provost Samantha, A. Cook, A. Dodson, A. Faudree, J. Greenhouse, S. Kar, L. Pessaint, M. Porto, K. Steingrebe, D. Valentin, S. Zouantcha, W. Bock Kevin, M. Minai, M. Nagata Bianca, R. van de Wetering, S. Boyoglu-Barnum, K. Leung, W. Shi, S. Yang Eun, Y. Zhang, M. Todd John-Paul, L. Wang, S. Alvarado Gabriela, H. Andersen, E. Foulds Kathryn, K. Edwards Darin, R. Mascola John, N. Moore Ian, G. Lewis Mark, A. Carfi, D. Montefiori, S. Suthar Mehul, A. McDermott, M. Roederer, J. Sullivan Nancy, C. Douek Daniel, S. Graham Barney, and A. Seder Robert, Immune correlates of protection by mRNA-1273 vaccine against SARS-CoV-2 in nonhuman primates. Science, 2021. 373(6561): p. eabj0299.

20. Krammer, F. and V. Simon, Serology assays to manage COVID-19. Science, 2020. 368(6495): p. 1060–1061.

21. Schultz-Cherry, S., M.A. McGargill, P.G. Thomas, J.H. Estepp, A.H. Gaur, E.K. Allen, K.J. Allison, L. Tang, R.J. Webby, S.D. Cherry, C.-Y. Lin, T. Fabrizio, E.I. Tuomanen, J. Wolf, and S.i. team, Cross-reactive antibody response to mRNA SARS-CoV-2 vaccine after recent COVID-19-specific monoclonal antibody therapy. Open Forum Infectious Diseases, 2021.

22. Cohen, M.S., A. Nirula, M.J. Mulligan, R.M. Novak, M. Marovich, C. Yen, A. Stemer, S.M. Mayer, D. Wohl, B. Brengle, B.T. Montague, I. Frank, R.J. McCulloh, C.J. Fichtenbaum, B. Lipson, N. Gabra, J.A. Ramirez, C. Thai, W. Chege, M.M. Gomez Lorenzo, N. Sista, J. Farrior, M.E. Clement, E.R. Brown, K.L. Custer, J. Van Naarden, A.C. Adams, A.E. Schade, M.C. Dabora, J. Knorr, K.L. Price, J. Sabo, J.L. Tuttle, P. Klekotka, L. Shen, D.M. Skovronsky, and B.-. Investigators, Effect of Bamlanivimab vs Placebo on Incidence of COVID-19 Among Residents and Staff of Skilled Nursing and Assisted Living Facilities: A Randomized Clinical Trial. JAMA, 2021.

23. Centers for Disease Control and Prevention, When You’ve Been Fully Vaccinated, Department of Health and Human Services, Editor. 2021: https://www.cdc.gov/coronavirus/2019-ncov/vaccines/fully-vaccinated.html.

24. Chigutsa, E., L. O’Brien, L. Ferguson-Sells, A. Long, and J. Chien, Population Pharmacokinetics and Pharmacodynamics of the Neutralizing Antibodies Bamlanivimab and Etesevimab in Patients With Mild to Moderate COVID-19 Infection. Clin Pharmacol Ther, 2021. 110(5): p. 1302–1310.

25. US Food and Drug Administration, Fact Sheet for Healthcare Providers Administering Vaccine - Emergency Use Authorization of the Pfizer-BioNTech Vaccine to Prevent Coronavirus Disease 2019 (COVID-19). 2021, Accessed 27 September 2021.: https://www.fda.gov/media/144413/download.

26. US Food and Drug Administration, Fact Sheet for Healthcare Providers Administering Vaccine (Vaccination Providers) - Emergency Use Authorization of the Moderna COVID-19 Vaccine to Prevent Coronavirus 2019 (COVID-19). 2021, Accessed 27 September 2021: https://www.fda.gov/media/144637/download.

27. Jones, B.E., P.L. Brown-Augsburger, K.S. Corbett, K. Westendorf, J. Davies, T.P. Cujec, C.M. Wiethoff, J.L. Blackbourne, B.A. Heinz, D. Foster, R.E. Higgs, D. Balasubramaniam, L. Wang, R. Bidshahri, L. Kraft, Y. Hwang, S. Žentelis, K.R. Jepson, R. Goya, M.A. Smith, D.W. Collins, S.J. Hinshaw, S.A. Tycho, D. Pellacani, P. Xiang, K. Muthuraman, S. Sobhanifar, M.H. Piper, F.J. Triana, J. Hendle, A. Pustilnik, A.C. Adams, S.J. Berens, R.S. Baric, D.R. Martinez, R.W. Cross, T.W. Geisbert, V. Borisevich, O. Abiona, H.M. Belli, M. de Vries, A. Mohamed, M. Dittmann, M. Samanovic, M.J. Mulligan, J.A. Goldsmith, C.-L. Hsieh, N.V. Johnson, D. Wrapp, J.S. McLellan, B.C. Barnhart, B.S. Graham, J.R. Mascola, C.L. Hansen, and E. Falconer, The neutralizing antibody, LY-CoV555, protects against SARS-CoV-2 infection in non-human primates Science Translational Medicine, 2021. eabf1906.

28. Shi, R., C. Shan, X. Duan, Z. Chen, P. Liu, J. Song, T. Song, X. Bi, C. Han, L. Wu, G. Gao, X. Hu, Y. Zhang, Z. Tong, W. Huang, W.J. Liu, G. Wu, B. Zhang, L. Wang, J. Qi, H. Feng, F.-S. Wang, Q. Wang, G.F. Gao, Z. Yuan, and J. Yan, A human neutralizing antibody targets the receptor-binding site of SARS-CoV-2. Nature, 2020. 584(7819): p. 120–124.

29. Starr, T.N., A.J. Greaney, A.S. Dingens, and J.D. Bloom, Complete map of SARS-CoV-2 RBD mutations that escape the monoclonal antibody LY-CoV555 and its cocktail with LY-CoV016. Cell Reports Medicine, 2021. 2(4): p. 100255.

30. Hoffmann, M., P. Arora, R. Groß, A. Seidel, B.F. Hörnich, A.S. Hahn, N. Krüger, L. Graichen, H. Hofmann-Winkler, and A. Kempf, SARS-CoV-2 variants B. 1.351 and P. 1 escape from neutralizing antibodies. Cell, 2021. 184(9): p. 2384-2393.e12.

31. Khoury, D.S., D. Cromer, A. Reynaldi, T.E. Schlub, A.K. Wheatley, J.A. Juno, K. Subbarao, S.J. Kent, J.A. Triccas, and M.P. Davenport, Neutralizing antibody levels are highly predictive of immune protection from symptomatic SARS-CoV-2 infection. Nature Medicine, 2021. 27(7): p. 1205–1211.

32. Steensels, D., N. Pierlet, J. Penders, D. Mesotten, and L. Heylen, Comparison of SARS-CoV-2 Antibody Response Following Vaccination With BNT162b2 and mRNA-1273. JAMA, 2021.

33. Baden, L.R., H.M. El Sahly, B. Essink, K. Kotloff, S. Frey, R. Novak, D. Diemert, S.A. Spector, N. Rouphael, C.B. Creech, J. McGettigan, S. Khetan, N. Segall, J. Solis, A. Brosz, C. Fierro, H. Schwartz, K. Neuzil, L. Corey, P. Gilbert, H. Janes, D. Follmann, M. Marovich, J. Mascola, L. Polakowski, J. Ledgerwood, B.S. Graham, H. Bennett, R. Pajon, C. Knightly, B. Leav, W. Deng, H. Zhou, S. Han, M. Ivarsson, J. Miller, and T. Zaks, Efficacy and Safety of the mRNA-1273 SARS-CoV-2 Vaccine. N Engl J Med, 2021. 384(5): p. 403–416.

34. Polack, F.P., S.J. Thomas, N. Kitchin, J. Absalon, A. Gurtman, S. Lockhart, J.L. Perez, G. Pérez Marc, E.D. Moreira, C. Zerbini, R. Bailey, K.A. Swanson, S. Roychoudhury, K. Koury, P. Li, W.V. Kalina, D. Cooper, R.W. Frenck, L.L. Hammitt, Ö. Türeci, H. Nell, A. Schaefer, S. Ünal, D.B. Tresnan, S. Mather, P.R. Dormitzer, U. Şahin, K.U. Jansen, and W.C. Gruber, Safety and Efficacy of the BNT162b2 mRNA Covid-19 Vaccine. New England Journal of Medicine, 2020. 383(27): p. 2603–2615.

35. Lustig, Y., E. Sapir, G. Regev-Yochay, C. Cohen, R. Fluss, L. Olmer, V. Indenbaum, M. Mandelboim, R. Doolman, S. Amit, E. Mendelson, A. Ziv, A. Huppert, C. Rubin, L. Freedman, and Y. Kreiss, BNT162b2 COVID-19 vaccine and correlates of humoral immune responses and dynamics: a prospective, single-centre, longitudinal cohort study in health-care workers. The Lancet. Respiratory medicine, 2021. 9(9): p. 999–1009.

36. Centers for Disease Control and Prevention, People with certain medical conditions. Certain medical condititions and risk for severe COVID-19 illness. November 2, 2020; Accessed September 23, 2021: https://www.cdc.gov/coronavirus/2019-ncov/need-extra-precautions/people-with-medical-conditions.html.

37. Iwasaki, A. and Y. Yang, The potential danger of suboptimal antibody responses in COVID-19. Nature Reviews Immunology, 2020. 20(6): p. 339–341.

38. Taylor, P.C., A.C. Adams, M.M. Hufford, I. de la Torre, K. Winthrop, and R.L. Gottlieb, Neutralizing monoclonal antibodies for treatment of COVID-19. Nature Reviews Immunology, 2021.

39. Müller, L., M. Andrée, W. Moskorz, I. Drexler, L. Walotka, R. Grothmann, J. Ptok, J. Hillebrandt, A. Ritchie, D. Rabl, P.N. Ostermann, R. Robitzsch, S. Hauka, A. Walker, C. Menne, R. Grutza, J. Timm, O. Adams, and H. Schaal, Age-dependent immune response to the Biontech/Pfizer BNT162b2 COVID-19 vaccination. Clin Infect Dis, 2021.

40. Bian, L., F. Gao, J. Zhang, Q. He, Q. Mao, M. Xu, and Z. Liang, Effects of SARS-CoV-2 variants on vaccine efficacy and response strategies. Expert Rev Vaccines, 2021. 20(4): p. 365–373.

41. Mlcochova, P., S.A. Kemp, M.S. Dhar, G. Papa, B. Meng, I.A.T.M. Ferreira, R. Datir, D.A. Collier, A. Albecka, S. Singh, R. Pandey, J. Brown, J. Zhou, N. Goonawardane, S. Mishra, C. Whittaker, T. Mellan, R. Marwal, M. Datta, S. Sengupta, K. Ponnusamy, V.S. Radhakrishnan, A. Abdullahi, O. Charles, P. Chattopadhyay, P. Devi, D. Caputo, T. Peacock, C. Wattal, N. Goel, A. Satwik, R. Vaishya, M. Agarwal, A. Mavousian, J.H. Lee, J. Bassi, C. Silacci-Fegni, C. Saliba, D. Pinto, T. Irie, I. Yoshida, W.L. Hamilton, K. Sato, S. Bhatt, S. Flaxman, L.C. James, D. Corti, L. Piccoli, W.S. Barclay, P. Rakshit, A. Agrawal, and R.K. Gupta, SARS-CoV-2 B.1.617.2 Delta variant replication and immune evasion. Nature, 2021.

42. Collier, D.A., A. De Marco, I.A.T.M. Ferreira, B. Meng, R.P. Datir, A.C. Walls, S.A. Kemp, J. Bassi, D. Pinto, C. Silacci-Fregni, S. Bianchi, M.A. Tortorici, J. Bowen, K. Culap, S. Jaconi, E. Cameroni, G. Snell, M.S. Pizzuto, A.F. Pellanda, C. Garzoni, A. Riva, N. Kingston, A. Elmer, B. Graves, L.E. McCoy, K.G.C. Smith, J.R. Bradley, N. Temperton, L. Ceron-Gutierrez, G. Barcenas-Morales, W. Harvey, H.W. Virgin, A. Lanzavecchia, L. Piccoli, R. Doffinger, M. Wills, D. Veeler, D. Corti, and R.K. Gupta, Sensitivity of SARS-CoV-2 B.1.1.7 to mRNA vaccine-elicited antibodies. Nature, 2021. 593(7857): p. 136–141.

43. Weinreich, D.M., S. Sivapalasingam, T. Norton, S. Ali, H. Gao, R. Bhore, B.J. Musser, Y. Soo, D. Rofail, J. Im, C. Perry, C. Pan, R. Hosain, A. Mahmood, J.D. Davis, K.C. Turner, A.T. Hooper, J.D. Hamilton, A. Baum, C.A. Kyratsous, Y. Kim, A. Cook, W. Kampman, A. Kohli, Y. Sachdeva, X. Graber, B. Kowal, T. DiCioccio, N. Stahl, L. Lipsich, N. Braunstein, G. Herman, and G.D. Yancopoulos, REGN-COV2, a Neutralizing Antibody Cocktail, in Outpatients with Covid-19. New England Journal of Medicine, 2020. 384(3): p. 238–251.

44. Zhang, L., J. Poorbaugh, M. Dougan, P. Chen, R.L. Gottlieb, G. Huhn, S. Beasley, M. Daniels, T. Ngoc Vy Trinh, M. Crisp, J.J. Freitas, P. Vaillancourt, D.R. Patel, A. Nirula, N. Kallewaard, R.E. Higgs, and R.J. Benschop, Endogenous antibody responses to SARS-CoV-2 in patients with mild or moderate COVID-19 who received bamlanivimab alone or bamlanivimab and etesevimab together Frontiers in Immunology, 2021. 12.

45. Baden, L.R., H.M. El Sahly, B. Essink, K. Kotloff, S. Frey, R. Novak, D. Diemert, S.A. Spector, N. Rouphael, C.B. Creech, J. McGettigan, S. Khetan, N. Segall, J. Solis, A. Brosz, C. Fierro, H. Schwartz, K. Neuzil, L. Corey, P. Gilbert, H. Janes, D. Follmann, M. Marovich, J. Mascola, L. Polakowski, J. Ledgerwood, B.S. Graham, H. Bennett, R. Pajon, C. Knightly, B. Leav, W. Deng, H. Zhou, S. Han, M. Ivarsson, J. Miller, and T. Zaks, Efficacy and Safety of the mRNA-1273 SARS-CoV-2 Vaccine. New England Journal of Medicine, 2020. 384(5): p. 403–416.

46. Elshal, M.F. and J.P. McCoy, Multiplex bead array assays: performance evaluation and comparison of sensitivity to ELISA. Methods, 2006. 38(4): p. 317–323.

47. Wang, P., M.S. Nair, L. Liu, S. Iketani, Y. Luo, Y. Guo, M. Wang, J. Yu, B. Zhang, P.D. Kwong, B.S. Graham, J.R. Mascola, J.Y. Chang, M.T. Yin, M. Sobieszczyk, C.A. Kyratsous, L. Shapiro, Z. Sheng, Y. Huang, and D.D. Ho, Antibody Resistance of SARS-CoV-2 Variants B.1.351 and B.1.1.7. bioRxiv, 2021: p. 2021.01.25.428137.

48. Nie, J., Q. Li, J. Wu, C. Zhao, H. Hao, H. Liu, L. Zhang, L. Nie, H. Qin, and M. Wang, Establishment and validation of a pseudovirus neutralization assay for SARS-CoV-2. Emerging microbes & infections, 2020. 9(1): p. 680–686.

49. Dogan, M., L. Kozhaya, L. Placek, C. Gunter, M. Yigit, R. Hardy, M. Plassmeyer, P. Coatney, K. Lillard, and Z. Bukhari, SARS-CoV-2 specific antibody and neutralization assays reveal the wide range of the humoral immune response to virus. Communications Biology, 2021. 4(1): p. 1–13.

50. Shankar, G., S. Arkin, L. Cocea, V. Devanarayan, S. Kirshner, A. Kromminga, V. Quarmby, S. Richards, C.K. Schneider, M. Subramanyam, S. Swanson, D. Verthelyi, and S. Yim, Assessment and Reporting of the Clinical Immunogenicity of Therapeutic Proteins and Peptides—Harmonized Terminology and Tactical Recommendations. The AAPS Journal, 2014. 16(4): p. 658–673.

51. Team, R.C., R: A language and environment for statistical computing. R Foundation for Statistical Computing. 2017.

52. Eastwood, B.J., M.W. Farmen, P.W. Iversen, T.J. Craft, J.K. Smallwood, K.E. Garbison, N.W. Delapp, and G.F. Smith, The Minimum Significant Ratio: A Statistical Parameter to Characterize the Reproducibility of Potency Estimates from Concentration-Response Assays and Estimation by Replicate-Experiment Studies. Journal of Biomolecular Screening, 2006. 11(3): p. 253–261.

53. Haas JV E.B., Iversen PW, Devanarayan, V., Weidner, J.R., Minimum Significant Ratio - A statistic to assess assay variability, in Assay Guidance Manual, G.A. Markossian S, Brimacombe K., Editor. 2017: https://www.ncbi.nlm.nih.gov/books/NBK169432/.

