## Supplementary Materials for "The effect of anti-SARS-CoV-2 monoclonal antibody, bamlanivimab, on endogenous immune response to COVID-19 vaccination"

This file includes:

Supplementary Text

Figs. S1 to S5

**Supplementary Text:**

*Endogenous antibody response against SARS-CoV-2 beta variant*

The immune response of participants was also evaluated against the SARS-CoV-2 beta variant (B.1.351), which bamlanivimab also does not significantly bind (Table 2) *(11)*. Compared with placebo, treatment with bamlanivimab resulted in a 1.7-fold lower titer against the beta variant (fig. S1a). These antibody titer data were grouped into participants who were either staff or residents (fig. S1b) and participants who received either SpikeVax or Comirnaty (fig. S3c). Mirroring the results shown in Fig. 2a, antibody titers from staff were 2.9-fold (p<0.001) higher than titers from residents against the beta variant (fig. S1b). The effect of bamlanivimab on vaccine-induced antibody titer against the beta variant (p=0.326) was similar for both residents and staff. There was also no significant difference in antibody titers for participants who received either SpikeVax or Comirnaty against the beta variant (p=0.842) (fig. S3c). For participants who received either vaccine, the effect of bamlanivimab on the vaccine-induced antibody titer against the beta variant was also not significantly different (p=0.77).

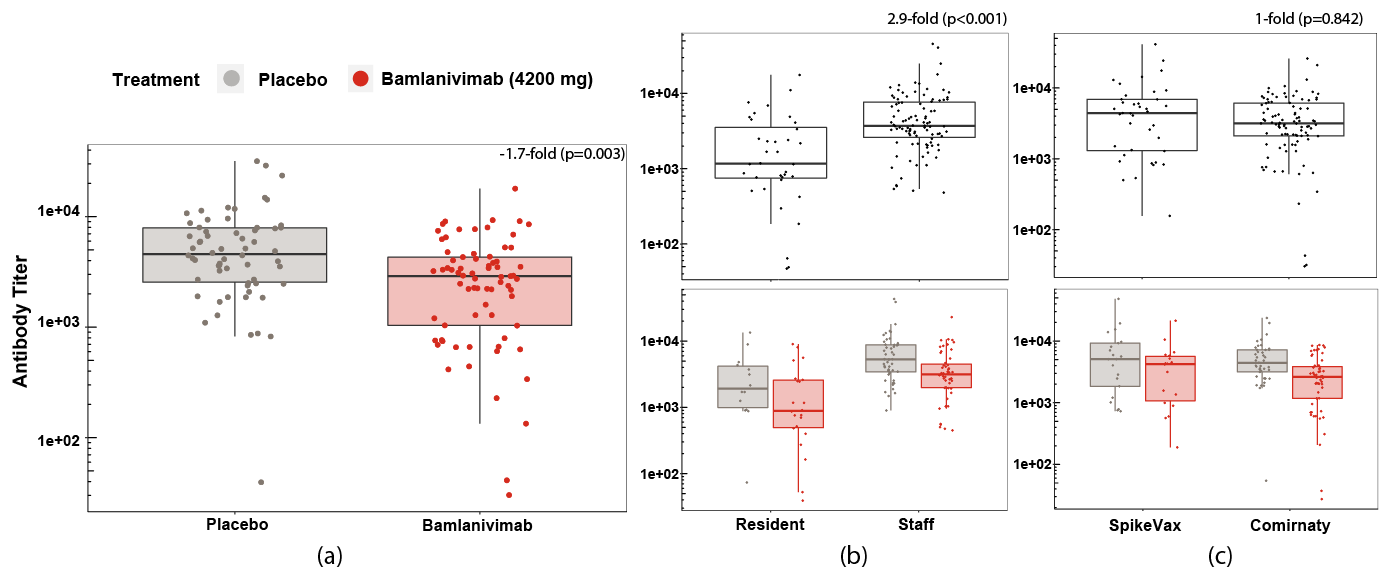

**Fig. S1:** Antibody titers against SARS-CoV-2 beta variant (B.1.351) for fully vaccinated participants who (a) previously received either bamlanivimab or placebo infusion (b) who were resident of staff and were subsequently fully vaccinated (SpikeVax or Comirnaty) against COVID-19. Antibody titers were rescaled after adjusting for covariates. Boxes and horizontal bars denote the interquartile range (IQR) and the median titer, respectively. Length of whiskers corresponds to 1.5 times the IQR.

*High-risk participants also mount robust antibody response to full COVID-19 vaccination following anti-SARS-CoV-2 mAb infusion*

Since the staff (N=99) included participants who were at high-risk of developing severe COVID-19 disease, an additional analysis compared the immune responses of high-risk staff participants (N=45), with non-high-risk staff participants (N=54). Of these 45 high-risk staff participants, 13% were over 65 years old and at high-risk by pre-specified definition *(13)*.

The intention of this comparison was to ascertain, firstly, whether antibody titers following full vaccination differs between these risk groups and secondly, whether bamlanivimab infusion disparately affected these risk groups. Antibody titers for non-high-risk staff and high-risk staff were similar against both E484Q and spike-NTD (p=0.594 and p=0.348, respectively) (fig. S2). The effect of bamlanivimab on vaccine-induced antibody titer against Spike-RBD-E484Q resulted in a significantly lower titer (-1.8-fold, p=0.037) for high-risk staff compared with non-high-risk staff (fig. S2a). However, this effect was not observed against spike-NTD, where the effect of bamlanivimab was similar for the two groups (p=0.249) (fig S2b).

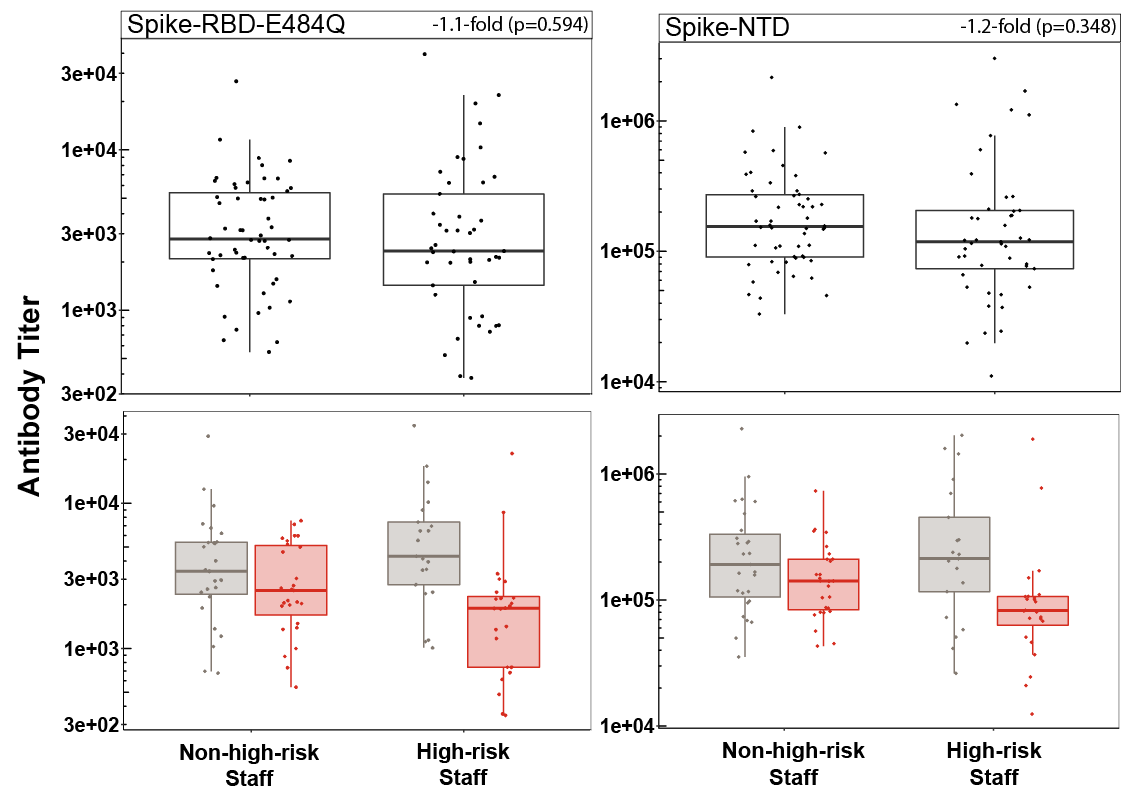

**Fig. S2:** Antibody titers against (a) Spike-RBD-E484Q and (b) spike-NTD comparing samples from fully vaccinated staff participants who were non-high-risk or at high-risk of developing severe COVID-19 disease (top) and further grouped by those who received placebo or bamlanivimab prior to vaccination (bottom). Antibody titers were rescaled after adjusting for covariates. Boxes and horizontal bars denote the interquartile range (IQR) and the median of reciprocal IC50, respectively. Length of whiskers corresponds to 1.5 times the IQR.

The antibody potency data was also compared for the staff to ascertain differences between the risk groups. Antibody potency was similar for non-high-risk and high-risk staff as measured as ACE2 binding inhibition potency (p=0.441) (fig. S3a), as well as pseudovirus neutralization potency (p=0.34) (fig. S3b). The effect of bamlanivimab was also similar for non-high-risk and high-risk staff as measured with ACE2 binding inhibition assay (p=0.084) and with the pseudovirus neutralization assay against spike-E484Q (p=0.085).

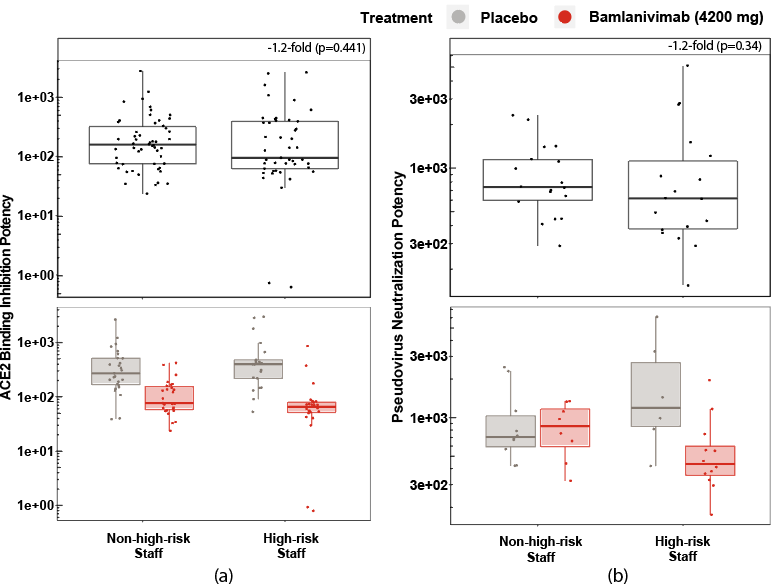

**Fig. S3:** Antibody potency (1/IC50) as measured using (a) ACE2 binding inhibition potency and (b) Spike-E484Q pseudovirus neutralization for fully vaccinated staff participants who were categorized as non-high risk or high risk (top) and further grouped by those who received placebo or bamlanivimab prior to vaccination. Potency measured as 1/IC50 and adjusted for covariates. Boxes and horizontal bars denote the interquartile range (IQR) and the median reciprocal IC50, respectively. Length of whiskers corresponds to 1.5 times the IQR.

*Neutralization potency against SARS-CoV-2 beta variant pseudovirus*

To corroborate the neutralization potency data against Spike-E484Q pseudovirus, the neutralization potency was also evaluated against the SARS-CoV-2 beta variant pseudovirus for the same subset of participants (N=49). There was also no statistically significant difference in the effect of bamlanivimab on pseudovirus neutralization potency against the beta variant compared with placebo (-1.2-fold, p=0.465) (fig. S4a). Furthermore, there was a significantly strong correlation (ρ=0.8, p<0.001) between the neutralization potency data against Spike-E484Q and the beta variant pseudoviruses (fig. S4b). These strong correlations also extended to pseudovirus data from participants who received either bamlanivimab or placebo (fig. S4b).

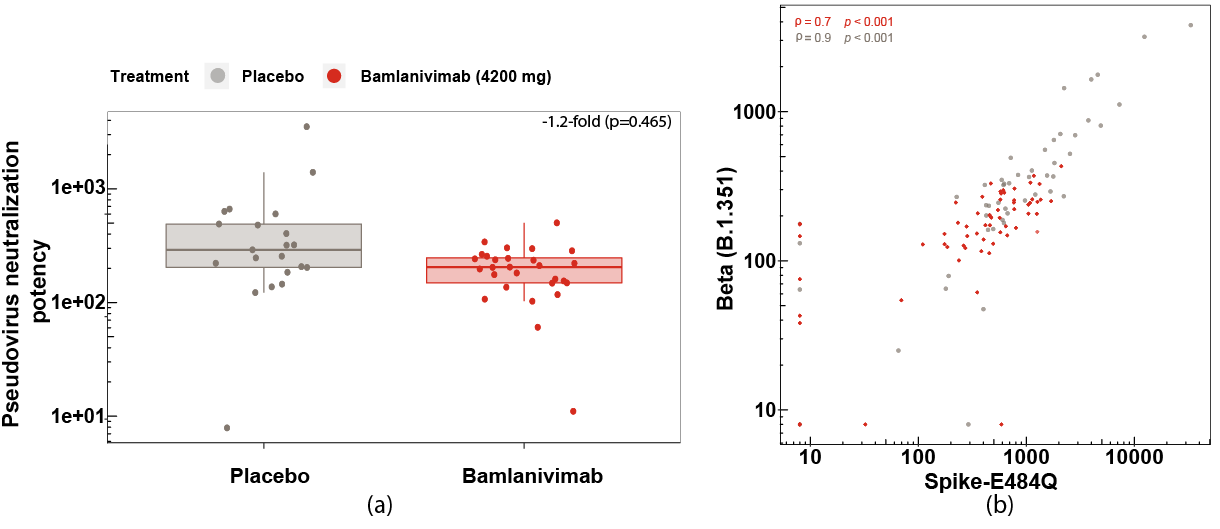

**Fig. S4:** (a) Neutralization potency (1/IC50) against SARS-CoV-2 beta variant (B.1.351) pseudovirus measured for samples collected from participants (N=49) who received placebo or bamlanivimab and were subsequently fully vaccinated. (b) Correlation plot of neutralization potency against beta variant and against Spike-E484Q. Pseudovirus neutralization potency measured as 1/IC50 and adjusted for T1 and T2 covariates. Boxes and horizontal bars denote the interquartile range (IQR) and the median reciprocal IC50, respectively. Length of whiskers corresponds to 1.5 times the IQR.

*Longitudinal observation of antibody titers against spike-NTD*

To visualize the longitudinal antibody responses to the COVID-19 vaccine, the antibody titers measured from samples (n=499) from all participants (N=135) were also evaluated against the spike-NTD. The same trends were observed as for the longitudinal antibody responses against Spike-RBD-E484Q (Fig. 6).

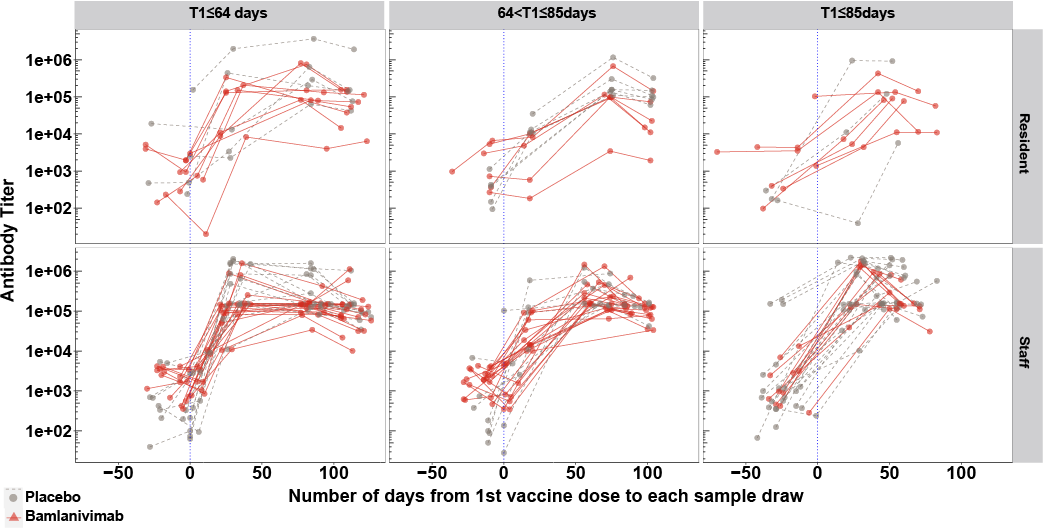

**Fig. S5:** Longitudinal antibody responses against spike-NTD protein arranged into three groups based on the interval (days) between bamlanivimab or placebo infusion and first vaccine dose, T1. Three columns (left to right) correspond to T1≤64 days, 64<T1≤85 days and T1>85 days, respectively. The vertical blue dotted line denotes the timepoint where participants receive the first dose of vaccine. Each line connects sample titers from a single participant. Top row shows antibody titers of participants who were residents and bottom row represents antibody titers of participants who were staff.

**Supplementary References**

1.	US Food and Drug Administration, *Fact sheet for health care providers: Emergency Use Authorization (EUA) of bamlanivimab* 2020: <https://www.fda.gov/media/143603/download>

2.	Chen, P., A. Nirula, B. Heller, R.L. Gottlieb, J. Boscia, J. Morris, G. Huhn, J. Cardona, B. Mocherla, V. Stosor, I. Shawa, A.C. Adams, J. Van Naarden, K.L. Custer, L. Shen, M. Durante, G. Oakley, A.E. Schade, J. Sabo, D.R. Patel, P. Klekotka, and D.M. Skovronsky, *SARS-CoV-2 Neutralizing Antibody LY-CoV555 in Outpatients with Covid-19.* New England Journal of Medicine, 2020. **384**(3): p. 229-237.
